# A non-coding single nucleotide polymorphism at 8q24 drives IDH1-mutant glioma formation

**DOI:** 10.1101/2022.03.08.22272044

**Authors:** Connor Yanchus, Kristen L. Drucker, Thomas M. Kollmeyer, Ricky Tsai, Minggao Liang, Lingyan Jiang, Judy Pawling, Asma Ali, Paul Decker, Matt Kosel, Arijit Panda, Ahmad Malik, Khalid N. Al-Zahrani, J. Javier Hernandez, Musaddeque Ahmed, Sampath Kumar Loganathan, Daniel Trcka, Antony Michaelraj, Jerome Fortin, Parisa Mazrooei, Lily Zhou, Andrew Elia, Mathieu Lupien, Housheng Hansen He, Liguo Wang, Alexej Abyzov, James W. Dennis, Michael D. Wilson, Jeffrey Wrana, Daniel Lachance, Margaret Wrensch, John Wiencke, Len A. Pennacchio, Diane E. Dickel, Axel Visel, Michael Taylor, Gelareh Zadeh, Peter Dirks, Jeanette E. Eckel-Passow, Tak Mak, Evgeny Kvon, Robert B. Jenkins, Daniel Schramek

**Affiliations:** Centre for Molecular and Systems Biology, Lunenfeld-Tanenbaum Research Institute, Mount Sinai Hospital; Toronto, Ontario, Canada; Department of Molecular Genetics, University of Toronto; Toronto, Ontario, Canada; Mayo Clinic, Rochester; Minnesota, USA; Hospital for Sick Children; Toronto, Ontario, Canada; Princess Margaret Cancer Centre, University Health Network; Toronto, Ontario, Canada; Department of Medical Biophysics, University of Toronto; Toronto, Ontario, Canada; Ontario Institute for Cancer Research; Toronto, Ontario, Canada; Department of Neurological Surgery; University of California, San Francisco; Environmental Genomics & System Biology Division, Lawrence Berkeley National Laboratory; Berkeley, USA; Comparative Biochemistry Program, University of California; Berkeley, USA; U.S. Department of Energy Joint Genome Institute; Berkeley, USA; School of Natural Sciences, University of California, Merced; Merced, USA; Department of Developmental & Cell Biology, University of California; Irvine, USA

## Abstract

Establishing causal links between inherited polymorphisms and cancer risk is challenging. Here, we focus on the single nucleotide polymorphism rs55705857 (A>G), which confers a 6-fold increased risk of IDH-mutant low-grade glioma (LGG) and is amongst the highest genetic associations with cancer. By fine-mapping the locus, we reveal that rs55705857 itself is the causal variant and is associated with molecular pathways that drive LGG. Mechanistically, we show that rs55705857 resides within a brain-specific enhancer, where the risk allele disrupts OCT2/4 binding, allowing increased interaction with the *Myc* promoter and increased *Myc* expression. To functionally test rs55705857, we generated an IDH1^R132H^-driven LGG mouse model and show that mutating the highly conserved, orthologous mouse rs55705857 locus dramatically accelerated tumor development from 463 to 172 days and increased penetrance from 30% to 75%. Overall, our work generates new LGG models and reveals mechanisms of the heritable predisposition to lethal glioma in ∼40% of LGG-patients.

## Introduction

The vast majority of cancer-related risk SNPs identified by genome-wide association studies (GWAS) are located in non-coding regulatory regions (*1, 2*). These GWAS tag-SNPs are usually themselves not the causal variant but are in linkage disequilibrium with one or more causative variants, which generally remain unknown. How such non-coding germline variants interact with acquired somatic mutations to facilitate cancer development is even less well understood. We previously identified several glioma susceptibility variants at 8q24.21, with rs55705857 A>G as the most significant SNP associated with a ∼6-fold increase in the relative risk of developing IDH-mutant low grade glioma (LGG) (*3-5*). This makes rs55705857 one of the highest reported inherited genetic associations with cancer, comparable with inherited *BRCA1* gene mutations and the risk of developing breast cancer or other familial glioma genes such as NF1/2, CDKN2A or p53 (**Fig. 1A)**. Interestingly, rs55705857 is not associated with the risk of *IDH*-WT glioma or other cancers, including *IDH*-mutant AML (*6-8*).

**Fig. 1.**
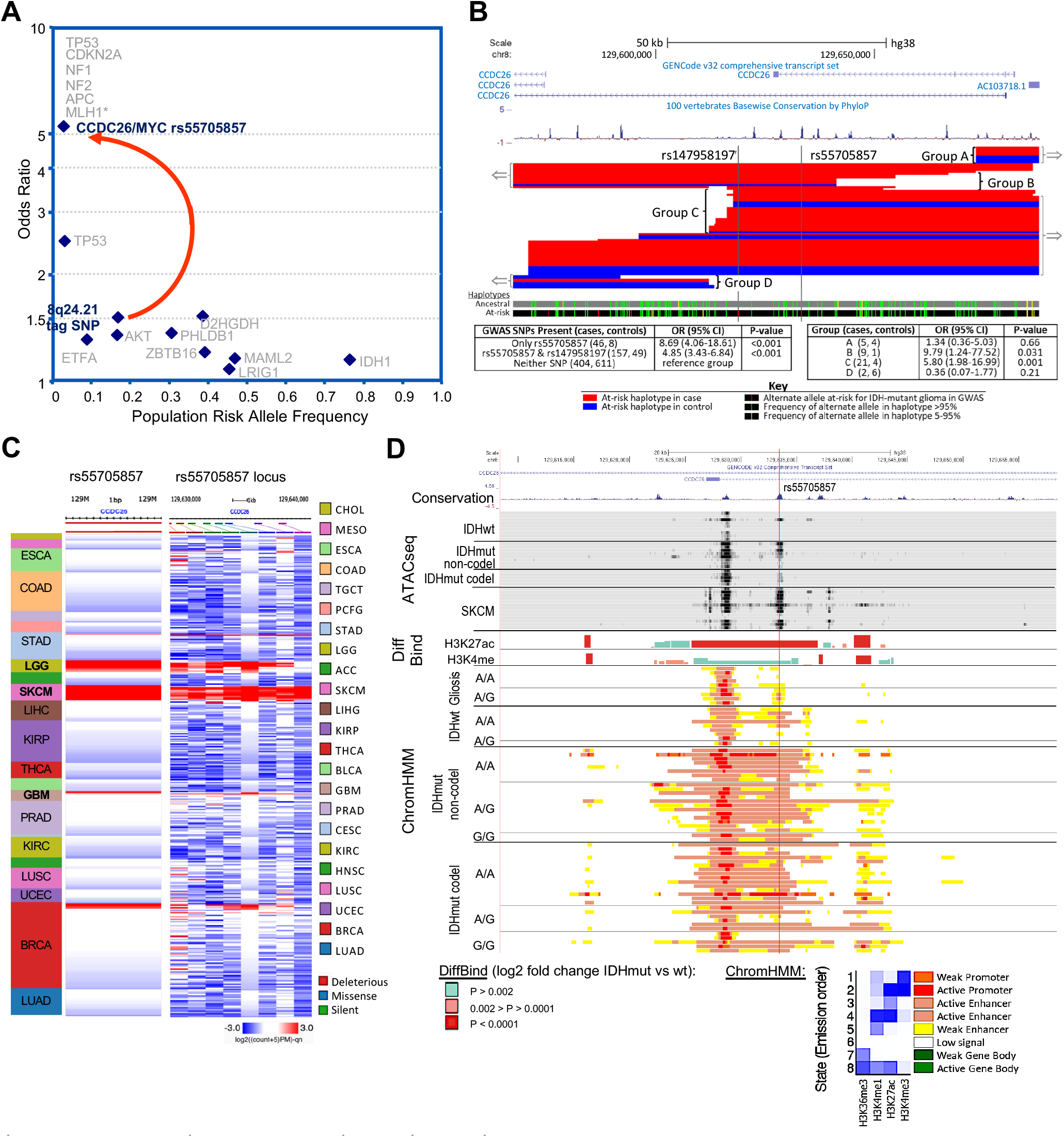
rs55705857 is the causal glioma risk variant at 8q24. (**A**) Fine-mapping of the 8q24 tag SNP allowed the discovery of a rs55705857 with much lower allele frequency and much higher effect size than the originally discovered tag SNP. Of the ten IDH mutant risk SNPs listed, rs55705857 has an odds ratio high enough to have an effect near that of familial inheritance glioma genes. *MLH1 is listed as a representative of the mismatch repair genes. (**B**) Fine-mapping of the minimal risk haplotype region surrounding the *IDH*-mutant glioma risk SNP rs55705857. Subjects heterozygous for the risk haplotype and with meiotic cross-overs disrupting the risk haplotype fall into 4 groups: two including the minimal overlap region (Groups B and C) and two not including the minimal overlap region (Groups A and D) (red =55 cases and blue= 22 controls). (**C**) ATAC-seq accessibility at genomic locus surrounding rs55705857 in 377 TCGA samples across 23 cancer types. (**D**) Gene transcripts, human/mouse conservation, and chromatin status surrounding rs55705857 are displayed. The red vertical line denotes the location of rs55705857. ATAC-seq data for the 8 *IDH*-WT and 13 *IDH*-mutant brain tumors and skin cutaneous melanoma (SKCM) are aligned with DiffBind log2-fold change for H3K27ac and H3K4me^1^ when comparing *IDH*-mutant vs. *IDH*-WT brain tumors. ChromHMM shows the predicted function of the genome surrounding rs55705857 based on the histone marks H3K36me3, H3K4me^1^, H3K27ac and H3K4me^3^ in *IDH*-WT and *IDH*-mutant brain tumors as well as non-tumor gliosis samples sorted by rs55705857 non-risk (A) and risk (G) alleles.

Lower grade glioma (LGG) are slow-growing brain tumors that eventually progress to aggressive glioblastoma. About 70% of LGG harbor transforming mutations in *IDH1* or *IDH2*, encoding isocitrate dehydrogenase (IDH) isozymes. Mutations mostly affect codon 132 of *IDH1* (R132H/C/S) or, less commonly, the homologous codon 172 of *IDH2* (R172K/W/G). While wild-type (WT) IDH isozymes metabolize isocitrate to α-ketoglutarate (αKG), mutant IDH reduce αKG to the oncometabolite R-2-hydroxygultarate (R-2HG), which alters the metabolic balance of affected cells (*9, 10*). Moreover, R-2HG is structurally similar to αKG and competitively inhibits αKG-dependent dioxygenases such as 5-methylcystosine (5mC) hydroxylases or histone lysine demethylases (KDMs). This gives rise to the characteristic LGG CpG island methylation phenotype and perturbs histone modifications and alters expression profiles of IDH-mutant gliomas (*11-13*). Based on their co-occurring genomic alterations, IDH-mutant LGG are subdivided into two types: molecular oligodendroglioma defined by co-deletion of chromosomal arms 1p and 19q (‘codel’) and *TERT* promoter mutations and the more aggressive molecular astrocytoma characterized by inactivation of *TP53* together with *ATRX* mutations (‘non-codel’) (*5, 14, 15*).

Here, we sought to reveal the molecular underpinnings for the specific and strong association between rs55705857 and *IDH*-mutant LGG as basis for understanding LGG initiation and heritable risk of developing LGG in ∼40% of LGG patients carrying the rs55705857 risk allele.

## Results

### Fine-mapping of inherited risk SNP variants at 8q24.21

To clarify whether rs55705857 G itself or other nearby SNPs were associated with LGG risk, we examined detailed haplotypes in genotyping data from 622 *IDH*-mutant LGG cases and 668 controls (*6, 7*). Identification of recombination events involving the risk haplotype allowed mapping the boundaries of the minimal region containing the causative variant (**Fig. 1B**). The minimal causative region contained only two loci, rs147958197 and rs55705857, that met genome-wide significance. However, some subjects with the rs55705857 G risk allele did not have the rs14758197 risk allele, but all subjects with the rs147958197 risk allele also had the rs55705857 G risk allele, suggesting that rs147958197 occurred on the haplotype already containing the rs55705857 G allele. Importantly, the odds ratios (OR) of the cases with only the rs55705857 risk allele and the cases carrying rs147958197 and rs55705857 risk alleles were not statistically different (p=0.19) (**Fig. 1B**). Results from sequencing six germline DNA samples containing a total of seven risk and five non-risk haplotypes did not identify any additional SNPs within the minimal causative region identifying rs55705857 as the likely causative 8q24.21 risk variant for IDH-mutant LGG.

### rs55705857 is located in an enhancer active in brain and LGG

rs55705857 resides in an intron of the lncRNA *CCDC26*, raising the possibility that this locus has a gene regulatory function. Mining Roadmap and ENCODE data revealed enrichment of histone modifications consistent with enhancer activity (H3K27ac, H3K4me^1^ and DNase I hypersensitivity) at the rs55705857 locus in neuronal and melanocyte linages but not in any other linages (**Fig. S1A**). Consistent with these data, examination of Assay for Transposase-Accessible Chromatin-sequencing (ATACseq) data from The Cancer Genome Atlas (TCGA) (*16*) revealed chromatin accessibility at rs55705857 almost exclusively in *IDH*-mutant LGG and cutaneous melanoma (**Fig. 1C, S2A**), suggesting that rs55705857 lies within an enhancer that is active in very selective cell lineages.

We next assessed epigenomic profiles of *IDH*-mutant human glioma. Chromatin-immunoprecipitation (ChIP)-seq revealed enrichment for the activating histone H3 lysine 27 acetylation (H3K27ac) and lysine 4 mono-methylation (H3K4me^1^) marks spanning rs55705857. This enhancer profile was more pronounced in *IDH*-mutant versus *IDH*-WT tumors, with 3.05- and 1.58-fold increased signals for H3K27ac and H3K4me^1^, respectively (diffbind; p=5.81×10^−7^ and p=2.31×10^−3^). Of note, active enhancer/promoter marks as inferred by the ChromHMM algorithm extended over 10 kb up- and downstream of rs55705857, which was not observed in either *IDH*-WT tumors or brain gliosis samples without tumors (**Fig. 1D and S1B**). However, there were no significant differences in H3K27ac and H3K4me^1^ in *IDH*-mutant tumors stratified by rs55705857 genotype (**Fig. 1D and S1B**). This suggests that *IDH* mutation, but not rs55705857 genotype, increases the enhancer activity of this locus in tumors.

### rs55705857 risk allele ‘G’ enhances an LGG-specific transcriptional profile

To delineate the functional impact of rs55705857, we performed expression quantitative trait loci (eQTL) analysis by correlating RNAseq transcriptional profiles with rs55705857 genotypes in 30 *IDH*-mutant codel, 29 *IDH*-mutant non-codel and 27 *IDH*-WT human gliomas. *PVT1* expression was significantly lower and *CCDC26* expression was significantly higher in *IDH*-mutant compared to *IDH-*WT tumors and *MYC* expression was significantly upregulated in all tumors compared to gliosis. However, the rs55705857 allele did not appear to alter expression of genes in the region, which was corroborated in TCGA data (**Table S1**). These results highlight that the transcriptional effect of IDH mutations is quite significant, while transcriptional impact of disease-associated polymorphisms may be subtle.

To delineate more subtle differences, we performed Gene Set Enrichment Analysis (GSEA) comparing *IDH*-mutant non-codel LGG tumors with gliosis and then within non-codel LGG stratified by rs55705857 genotype. Interestingly, both comparisons showed up-regulation of similar gene sets involved in epithelial to mesenchymal transition (EMT), IL-2 and IL-6 signaling, Inflammatory Response, Hypoxia, G2M checkpoints, p53 pathway, Interferon, as well as TNF signaling and a strong down-regulation of genes involved in oxidative phosphorylation (**Fig. S2B**). This suggests that rs55705857 risk allele might have an important functional role in augmenting the underlying biology of LGG. Given that the rs55705857 is in the ∼ 2Mb region that is regulating expression of the MYC oncogene in several cancers (*17*), we analyzed MYC gene sets, which showed enrichment in *IDH*-mutant non-codel tumors compared to gliosis. Interestingly, the rs55705857 risk allele is associated with upregulation of one MYC subset (MYC Targets V2) but downregulation of another (MYC Targets V1) (**Fig. S2B, Table S2**).

### rs55705857 risk allele ‘G’ increases and broadens enhancer activity in a mouse reporter assay

The extreme conservation of the non-risk ‘A’ allele at rs55705857 locus and its surrounding sequence across all mammalian species, including mice and even platypus (*4*) (**Fig. 1C**), prompted us to assess whether rs55705857 variants influence enhancer function *in vivo* in mice. First, we generated knock-in mice with site-specific integration of an enhancer construct comprised of the highly conserved 3225bp-long human fragment, with the rs55705857 non-risk A-allele (hs1709A) or risk G-allele (hs1709G) at the center of this fragment, followed by a minimal promoter and a *lacZ* reporter gene integrated into the *H11* safe harbor locus (**Fig. 2A**) (*18*). Both enhancer alleles were active in the cells of developing skin at embryonic day E11.5, consistent with a melanoblast staining pattern. Notably, the variant hs1709G had additional enhancer activity in the somite/rib area not observed for the hs1709A allele (**Fig. 2B and C**). At E14.5, both enhancer alleles became active in the neural tube, forebrain and the ribs. Strikingly, the risk-associated allele displayed an overall stronger enhancer activity in these structures and showed additional activity in the midbrain (**Fig. 2B and C**), indicating that the rs55705857 risk allele directly influences strength and tissue specificity of this developmental enhancer.

**Fig. 2.**
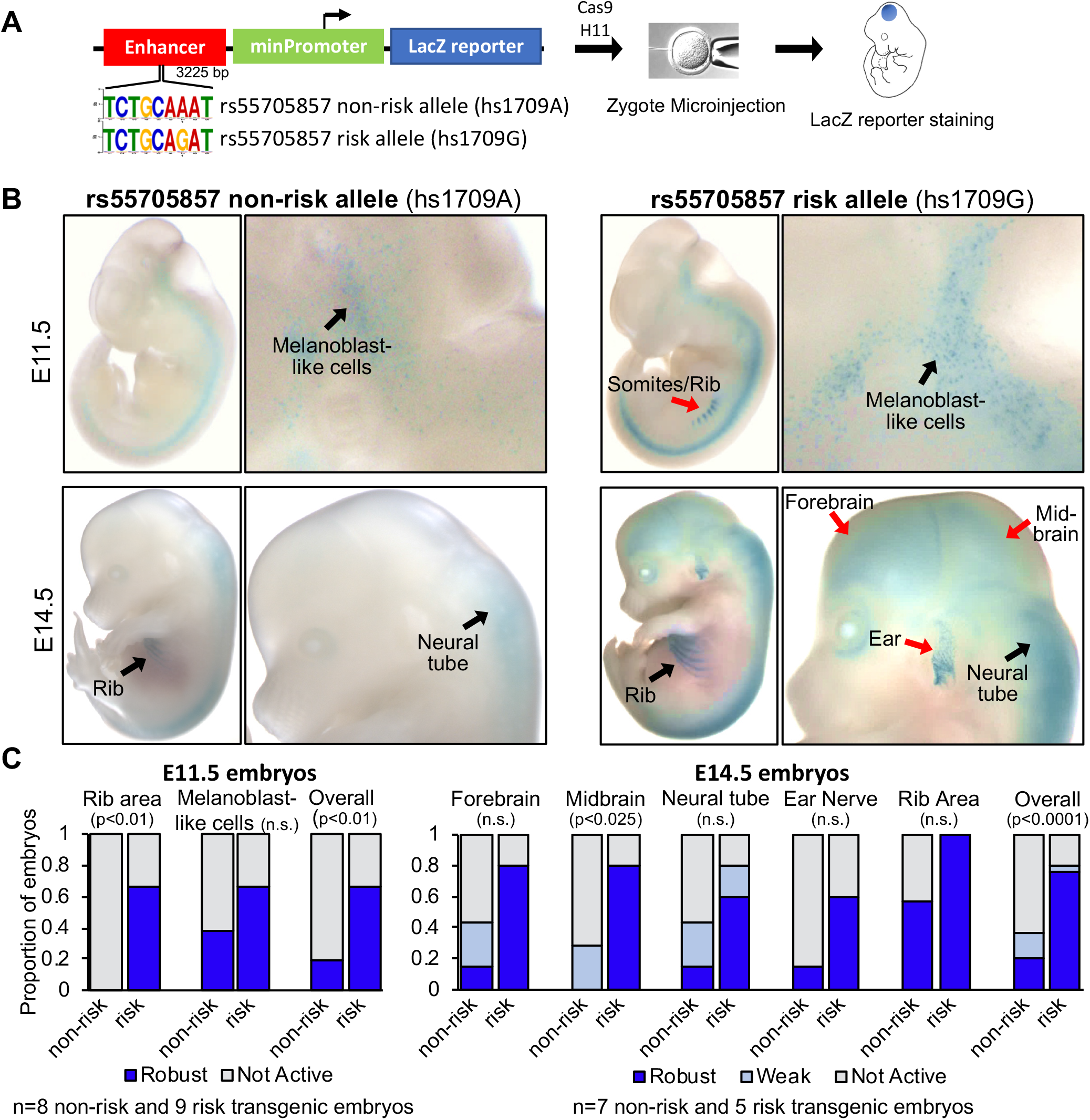
rs55705857 SNP resides in a brain-specific enhancer. (**A**) Schematic of rs55705857 enhancer reporter construct. (**B**) Representative whole mount images of LacZ-stained rs55705857 non-risk (left) and risk (right) enhancer reporter embryos. Black arrows denote LacZ staining found in both reporter mice while red arrows indicate LacZ staining only found in risk reporter mice. (**C**) Summary for enhancer activity of the non-risk and risk allele. (p-value by Fisher-Freeman-Halton Exact Test)

### Establishing a Mouse Model of *Idh1*^R132H^-mutant LGG

To determine how the rs55705857 locus effects gliomagenesis, we established an *IDH*-mutant LGG model using conditional *Idh1*^R132H^ knock-in mice (**Fig. 3A**) (*19*). As expected, transducing primary neural stem cells (NSC) from these *Idh1*^*LSL*-R132H/+^ mice with an adenovirus expressing Cre resulted in R-2HG accumulation and dramatically effected the tricarboxylic acid (TCA) cycle, the glycolysis and glutaminolysis pathways as well as the levels amino acids and nucleotides (**Fig. S3A-F**). We crossed the *Idh1*^*LSL*-R132H/+^ mouse to conditional *Trp53*^LSL-R270H/+^ mice, allowing for concomitant expression of p53^R270H^ (homologous to the human p53^R273H^). p53^R273H^ functions in a dominant negative manner, can have gain-of-function activity (*20*), and is the most prevalent p53 mutation found in human LGG (**Fig. S4A)**. To enable CRISPR/Cas9-mediated somatic mutagenesis of any other LGG-associated genes, we further crossed these mice to the LSL-Cas9-GFP mice. To induce the expression of IDH1^R132H^, p53^R270H^ and Cas9-GFP, we used stereotactic injections to deliver Cre-expressing lentiviral particles to the NSC residing in the lateral subventricular zone at postnatal day 0 (p0), which resulted in clonal induction of IDH1^R132H^ expression and accumulation of R-2HG (**Fig. S4B-E)**. Next, we assessed the knock-out efficiency of CRISPR/Cas9 using either a dual fluorescence-based reporter assay or targeting endogenous genes such as *Urod* or *Atrx*, revealing a knock-out efficiency of 60-85% (**Fig. S5A-E**).

**Fig. 3.**
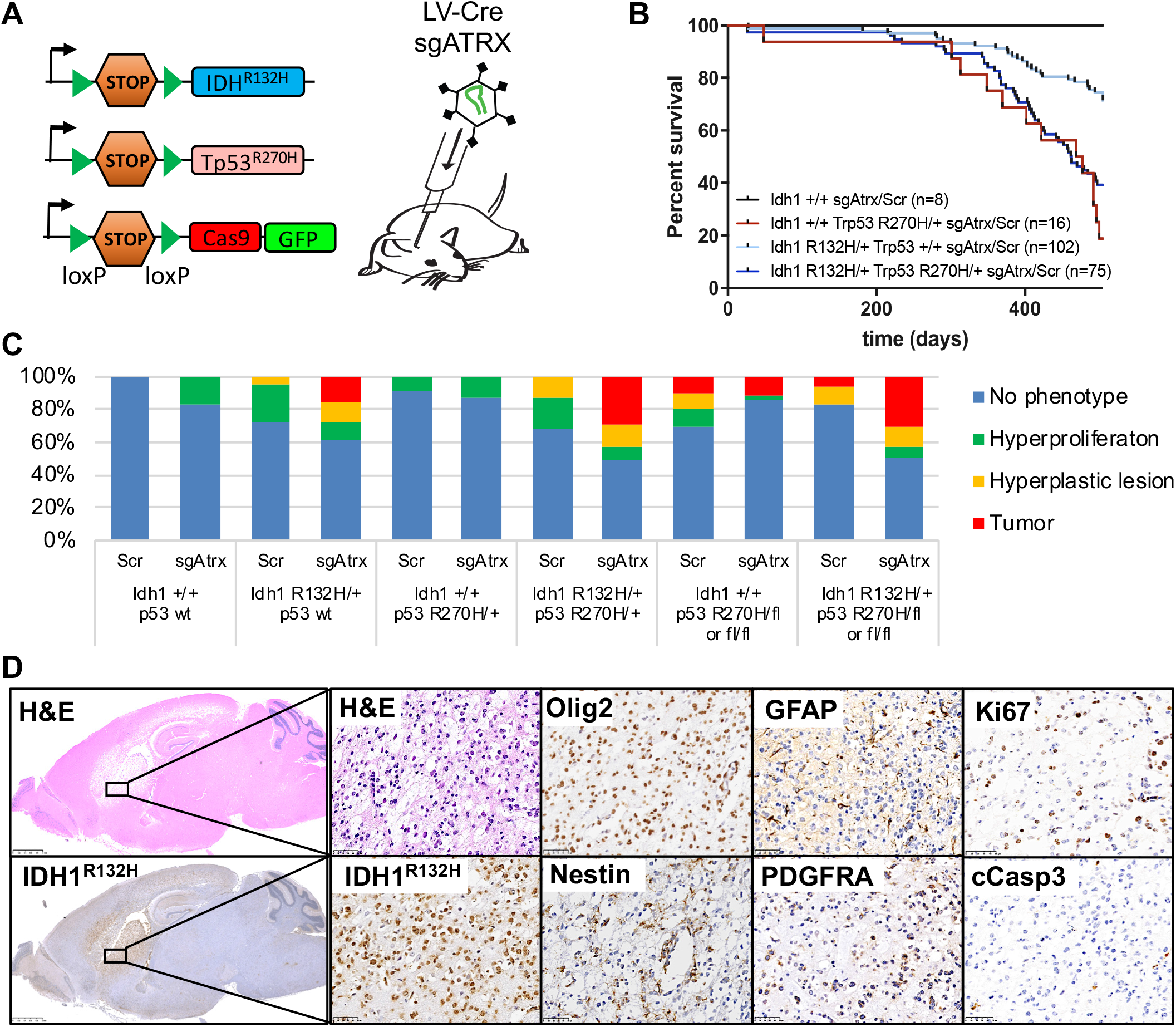
IDH1-mutant LGG mouse model. (**A**) Schematic of conditional alleles and CRISPR virus used to generate the LGG cohorts. (**B**) Survival of mice with the indicated genotype transduced with an sgRNA targeting *Atrx* or a scrambled control sgRNA (Scr). (n=201 mice; p < 0.0001, Log-rank (Mantel-Cox) test). (**C**) Bar graph indicating percentage of phenotypes found in mice from (B) with the indicated genotype. (**D**) Representative H&E and IHC staining of the same tumor region within a *Idh1*^R132H/+^;*Trp53*^R270H/+^;*Atrx*^-/-^;Cas9-GFP brain using indicated antibodies. Scale bars, 2.5mm (left) and 50µm (right).

Next, we generated cohorts of R26-Cas9-GFP mice with different combinations of *Idh1*^R132H^ and *p53* mutations and transduced them either with a LV-sg*Atrx*-Cre or a non-targeting, scrambled LV-sgScr-Cre virus. Starting at 301 days, we observed sarcomas and lymphomas in *Idh1*-wildtype *Trp53*^LSL-R270H/+^ mice necessitating euthanasia (**Fig. 3B)**. This is likely because the STOP cassette makes the *Trp53*^LSL-R270H/+^ mouse heterozygous for *Trp53*, which promotes spontaneous sarcoma and lymphoma development (*21, 22*). None of these *Idh1*-wildtype *Trp53*^LSL-R270H/+^ mice developed brain tumors. In contrast, 20% of *Idh1*^R132H/+^;*Trp53*^+/+^ and 30% of *Idh1*^R132H/+^;*Trp53*^R270H/+^ mice transduced with sg*Atrx* developed brain tumors in the cerebral cortex, cerebral striatum or OB with a median survival of 463 days. An additional 13-14% of these mice exhibited hyperplastic lesions in the brain (**Fig. 3B-D, S6A and B)**. Of note, induction of IDH1^R132H^ alone or in combination with p53^R270H^ but without targeting *Atrx* did not initiate glioma formation over a 500 day period (**Fig. 3C)**, consistent with previous reports (*23, 24*).

Non-codel LGG is usually associated with loss-of-heterozygosity of chr17p encompassing the *TP53* locus, suggesting biallelic *TP53* inactivation (*23*). Therefore, we generated cohorts of *Idh1*^+/+^ and *Idh1*^R132H/+^ mice harboring either two *Trp53*^fl^ alleles (*Trp53*^fl/fl^) or one *Trp53*^*fl*^ and one *Trp53*^*LSL-R270H*^ allele (*Trp53*^LSL-R270H/fl^). About 10% of *Idh1*^+/+^*;Trp53*^***fl/fl***^ or *Idh1*^+/+^;*Trp53*^LSL-R270H/fl^ transduced with LV-sgScr-Cre or LV-sg*Atrx*-Cre developed brain tumors as expected for mice with bi-allelic *Trp53* mutations (*22*) (**Fig. 3C)**. Interestingly, with regards to IDH1^R132H^-driven tumorigenesis, we did not observe a difference in tumor prevalence between heterozygous p53^R270H^ (*Trp53*^R270H/+^), complete loss of p53 (*Trp53*^fl/fl^) or p53^R270H^ with loss of the WT p53 allele (*Trp53*^LSL-R270H/fl^). About 30% of mice in all those cohorts developed brain tumors with similar latency and histology when transduced with LV-sg*Atrx*-Cre, while most LV-sgScr-Cre transduced mice stayed tumor-free (**Fig. 3C and S6C)**. These data indicate that p53^R270H^ functions in a dominant negative manner without apparent gain-of-function effects in this mouse model and demonstrate that IDH1^R132H^ cooperates with *Atrx* and *Trp53* mutations in the development of LGG.

All tumors expressed IDH1^R132H^ and harbored cells positive for Ki67, Olig2, Nestin, GFAP, PDGFRA, and exhibited a well-differentiated fibrillary neoplastic histology and low apoptotic cell numbers, recapitulating histopathological and molecular hallmarks of human LGG (**Fig. 3D and S6A-C)**. Expression profiling followed by GSEA comparing *Idh1*^R132H^, *Trp53*^R270H^ and *Atrx* compound mutant tumors to WT brain parenchyma revealed differentially expressed gene sets specifically associated with EMT, IL-2, Hypoxia, G2M checkpoints, p53 pathway, Interferon, mTOR signaling, TNF signaling, Myc and OxPhos (**Fig. S7A**), reminiscent of human *IDH*-mutant non-codel LGG (**Fig. 1D**). Cluster analysis with human gliomas of similar subtype confirmed that the mouse tumors faithfully recapitulate the human disease (**Fig. S6B**).

### Disruption of rs55705857 Increases Penetrance and Decreases Latency of IDH1^R132H^-driven Glioma

To assess the pathologic potential of rs55705857, we generated two mouse strains to evaluate the role of the highly orthologous mouse rs55707857 locus in modulating gliomagenesis. One mouse line harbors an orthologous rs55707857 A>G substitution in conjunction with a 4bp indel destroying the PAM site (denoted rs557^A->G+4^) and another line harbors a 66bp deletion (denoted rs557^66bp^) spanning the murine rs55705857 locus (**Fig. S8A-C**). Both lines (collectively rs557^mut^ mice) were viable, fertile, and displayed no overt phenotype or abnormal brain histology, indicating that the mutating the murine rs55705857 locus did not influence development.

We crossed these rs557^mut^ strains with *Idh1*^LSL-R132H/+^;*Trp53*^fl/fl^;LSL-Cas9-GFP mice and injected them with LV-sgScr-Cre or LV-sg*Atrx*-Cre. Both rs557^mut^ lines exhibited significantly increased the penetrance and drastically decreased latency of tumor formation compared to rs557^+/+^ (**Fig. 4A and B)**. While only 5% of the rs557^mut^;*Idh1*^+/+^;*Trp53*^-/-^ animals injected with either sgScr or an sg*Atrx* developed brain tumors, 34% and 75% of rs557^mut^;*Idh1*^R132H/+^;*Trp53*^fl/fl^ animals injected with sgScr or sg*Atrx* developed brain tumors, respectively. The median survival of rs557^mut^;*Idh1*^R132H/+^;*Trp53*^fl/fl^ animals injected with sg*Atrx* was 172-days for rs557^A->G+4^ and 201-days for rs557^66bp^ compared to the median survival of 463 days in rs557 control mice. Tumor location and histopathology were not altered compared to rs557^wt^ (**Fig. 4C**). Moreover, orthoptic transplanting rs557^66bp^;*Idh1*^R132H/+^;*Trp53*^-/-^ tumor cells (RIP cells) into recipient mice resulted in the formation of lethal gliomas (**Fig. 4D and S7C**). Together, these data clearly demonstrate that disruption of the rs55705857 locus facilitates glioma development.

**Fig. 4.**
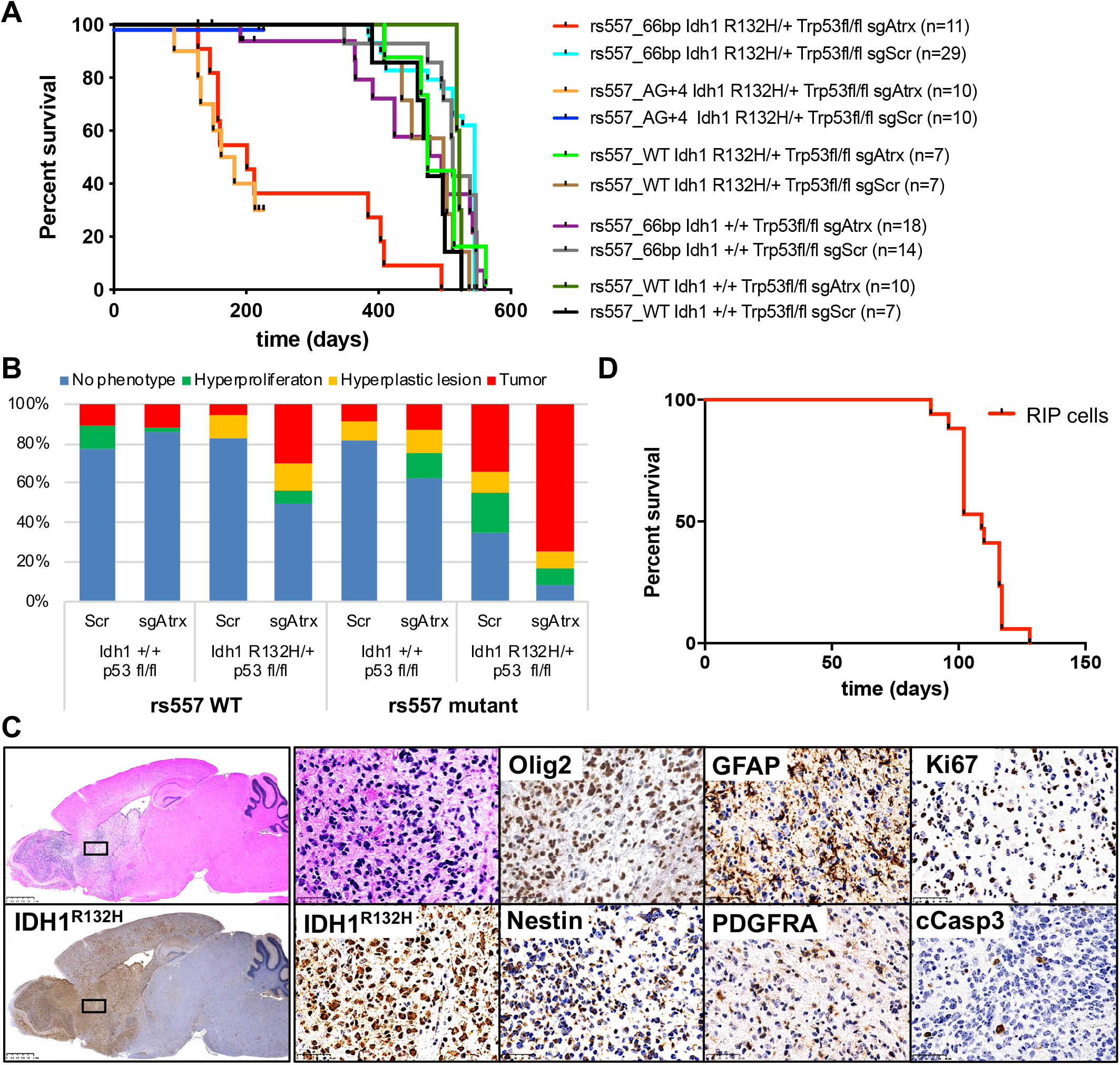
rs55705857 cooperates with IDH1, p53 and ATRX mutations. (**A**) Survival mice with the indicated genotype transduced with an sgRNA targeting *Atrx* or a scrambled control sgRNA (Scr). (n=123 mice; p-value < 0.0001, Log-rank (Mantel-Cox) test) (**B**) Bar graph indicating percentage of phenotypes found in mice from (A). (**C**) Representative H&E and IHC of the same tumor region within a rs557^66bp/+^;*Idh1*^R132H/+^;*Trp53*^fl/fl^;*Atrx*^-/-^; Cas9-GFP brain using indicated antibodies. Scale bars, 2.5mm (left) and 50µm (right). (**D**) Survival of Nod/Scid/γ mice intracranial injected with rs557^66bp/+^;*Idh1*^R132H/+^;*Trp53*^γ/ γ^;Cas9-GFP RIP cells. (n=17)

### The rs55705857 risk allele G disrupts an OCT transcription factor binding site

As SNPs in regulatory regions can modulate transcription factor binding, we performed motif analysis, which revealed that rs55705857 resides in an OCT2/4 binding motif (**Fig. 5A**). Notably, IntraGenomic Replicates (IGR) algorithm predicted the risk-associated G-allele to have a significantly lower binding intensity for OCT2/4 compared to the reference A-allele ∼1.8-fold (t-test -log10(p) = 3.09). In addition, the OCT2/4 motif is flanked by a highly conserved SOX2/4/9 and an ASCL1/2 motif (**Fig. 5B**), all of which play crucial roles in brain development (*25-28*) and gliomagenesis (*29-31*).

**Fig. 5.**
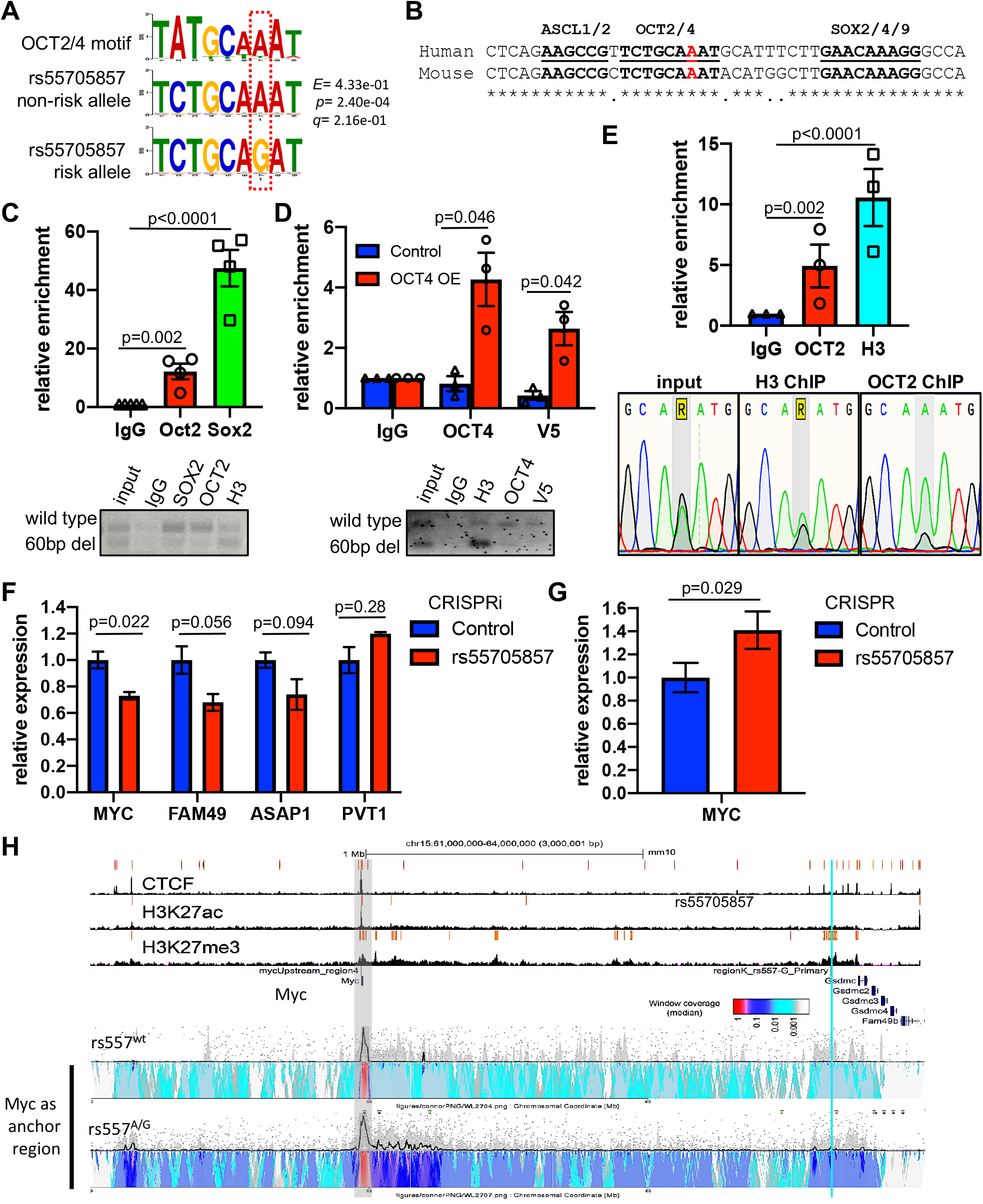
rs55705857 modulates OCT2 and OCT4 binding and regulates MYC expression. (**A**) The canonical OCT2/4 binding motif (top) and the rs55705857 non-risk ‘A’ allele (middle) and the rs55705857 risk ‘G’ allele (bottom) are shown. (**B**) Sequence alignment of the human rs55705857 and its orthologous mouse locus highlighting conserved binding motifs for ASCL1/2, OCT2/4, and SOX2/4/9. The non-risk rs55705857 A-allele is marked in red. * indicates conserved amino acids. (**C**) Enrichment of OCT2 and SOX2 at mouse rs55705857 locus. Top: ChIP-qPCR using rs557^66bp/+^, *Idh1*^R132H/+^; *Trp53*^γ/γ^;Cas9-GFP RIP cells (n = 4). IgG serves as a negative control and histone H3 as a positive control. (p-value by two-tailed t-test) Bottom: Representative gel electrophoresis analysis of PCR amplicons from IgG, SOX2, OCT2 and H3 ChIP. **(D)** Enrichment of OCT4 at the mouse rs55705857 locus. Top: ChIP-qPCR using rs557^66bp/+^, *Idh1*^R132H/+^; *Trp53*^γ*/*γ^;Cas9-GFP RIP cells transfected with an V5-tagged OCT4 performed using an OCT4 and an anti-V5 antibody. IgG serves as a negative control and histone H3 as a positive control. (p-value by two-tailed t-test) Bottom: Representative gel electrophoresis analysis of PCR amplicons from IgG, H3, OCT4 and V5 ChIP. (**E**)The risk allele G of rs55705857 disrupts OCT binding. Top: ChIP-qPCR showing enrichment of OCT2 at rs55705857 locus of human LGG cells heterozygous for the rs55705857 risk allele. IgG-IP serves as a negative control and histone H3 as a positive control. (p-value by two-tailed t-test) Bottom: Sanger sequencing chromatograms of the SNP region from input, histone H3 and OCT2 ChIPed DNA. (**F**) Expression of rs55705857 neighboring genes measured by real-time qPCR without or with silencing the rs55705857 locus using CRISPRi. (p-value by two-tailed t-test. (**G**) Relative expression of *Myc* measured by real-time qRT-PCR in rs557^66bp/+^ versus rs557-double mutant RIP cells. (p-value by two-tailed t-test) (**H**) Analysis of high frequency interacting regions at the *Myc* locus in rs55705857 WT versus A->G+4bp cells by 4C-seq. The heat map color-scale shows normalized median contact frequency. The black trendline shows the median contact frequency and the shaded grey area indicates the 20th to 80th percentiles. The light blue line marks the location of rs55705857, and shaded gray box marks the location of Myc.

To experimentally test whether the rs55705857 G allele affects OCT2/4 chromatin-binding, we performed ChIP in *Idh1*^R132H^-mutant RIP cells using OCT2, OCT4, SOX2, H3K4me^1^, and H3K27ac and antibodies followed by PCR amplification of the rs55705857 locus (ChIP-PCR). Both H3K4me^1^ and H3K27ac marked the murine rs55705857 locus (**Fig. S8D)**, reminiscent of the human LGG data (**Fig. 1C and S1B**). While OCT2, SOX2 and OCT4 were expressed at low levels in our murine tumors, reminiscent of their low-level expression in human LGG and GBMs (**Fig. S8E-F)**, OCT4 expression was lost upon culturing the tumor cells. Thus, we performed ChIP-PCR on endogenous OCT2 and SOX2 but had to exogenously express OCT4 (**Fig. S8G**). We found that OCT2, SOX2 and OCT4 bound preferentially to the murine rs55705857 WT allele compared to the mutant rs557^66bp^ allele (**Fig. 5C and D**). To extend these finding to humans, we performed OCT2 ChIP-PCR on human heterozygous rs55705857 A/G *IDH1*-mutant LGG cells. ChIP-PCR followed by sanger sequencing of the PCR amplicon revealed that OCT2 indeed preferentially binds the A allele (**Fig. 5E)**. Together, these data show that the rs55705857 SNP ‘G’ allele disrupts binding of OCT2/4.

### rs55705857 regulates Myc pathway and physically interacts with the *Myc* locus

To test whether the rs55705857 locus regulates expression of nearby genes, we first performed CRISPR interference (CRISPRi) targeting the rs55705857 locus in RIP cells, which led to reduced expression of *Myc* and other neighboring genes (**Fig. 5F**). Next, we established syngeneic RIP cells, where the WT allele was also mutated by CRISPR/Cas9 (**Fig. S7H)**. RIP cells with two mutant rs55705857 alleles compared to RIP cells harboring one WT rs55705857 allele exhibited modest but significant increased *Myc* expression (**Fig. 5G**), indicating that the rs55705857 WT ‘A’ allele functions to repress *Myc* expression.

Given the prominent role of MYC in carcinogenesis, we next investigated a potential interaction of rs55705857 with the *Myc* promoter using circular chromosome conformation capture assay (4C-seq). Supporting a potential role for rs55705857-G allele regulating *MYC* expression, 4C-seq experiments using rs557 ^A->G+4/+^ cells, but not rs557^WT^ cells, showed evidence of interactions with the rs55705857 locus (**Fig. 5H**). To extend these findings to humans, we analyzed Hi-C interaction data from healthy human hippocampus and dorsolateral prefrontal cortex (*32, 33*), which showed interactions of rs55705857 locus from both the rs55705857 and *MYC* perspective, including the *MYC* promoter and *PVT1* and several other loci between the two regions (**Fig. S9**). Together these data support a model where the rs55705857-G allele abrogates OCT2/4 binding within a conserved enhancer element allowing it to interact with *MYC* promoter and upregulate *MYC* expression.

## Discussion

By comprehensively profiling a large cohort of LGG, we found that rs55705857 itself is the causal risk variant and lies within a conserved OCT2/4 transcription factor binding motif within a brain-specific enhancer, which is hyperactivated in IDH-mutant LGGs. It is well known that 2-HG produced by mutant IDH competitively inhibits histone lysine demethylases such as KDM6A/B, resulting regional variation in histone modification; including areas of decreased and increased H3K27ac and H3K4me^1^ and enhancer activity (*11-13*). The region surrounding rs55705857 is clearly an area of increased enhancer activity. The hyperactive chromatin status combined with the tissue specificity of this enhancer thus explains the cooperativity between mutant IDH and rs55705857 and why rs55705857 is associated specifically with *IDH*-mutant glioma, but not other brain cancers.

Interestingly, we found that the rs55705857 locus functions as an enhancer not only in the brain but also in melanocytes. 5% of melanoma harbor *IDH1*^R132^ hotspot mutations. There is a well-documented increased risk of melanoma in glioma patients and this association is thought to be the result of common genetic predispositions. Germline deletion of the INK4 locus and alterations in telomere maintenance are associated with the melanoma-astrocytoma syndrome (*34-37*). It will be interesting to assess whether the rs55705857 G-risk allele also increases the susceptibility to melanoma.

Mechanistically, we show that the rs55705857 risk allele ‘G’ abrogates OCT2/4 binding to this enhancer, exhibits increased physical interactions with the *Myc* promoter and increased *Myc* transcription, indicating that OCT2/4 binding the non-risk rs55705857 locus restricts *MYC* expression. Apart from the well-known functions of OCT4 in activating transcription, OCT4 has been shown to act as a repressor of linage-specific transcription during early embryonic development (*38, 39*). In addition, OCT2 is a well-known transcriptional repressor and known to regulate neuronal differentiation (*40*). While the GSEA shows that rs55705857 enhances the expression of MYC targets, the possibility exists that rs55705857 additionally interacts with genes other than MYC in *cis* or *trans*, and acts through them in modulating tumor growth. In fact, the GSEA in human LGG demonstrates that the rs55705857 risk allele reinforces the biological pathways driving gliomagenesis.

To model IDH1-mutant glioma, we used conditional *Idh1*^R132H^ knock-in mice generated tumors by injecting Cre into newborn mice, which suggests that the initiation of human LGG can occur very early in life and is consistent with the diagnosis of *IDH*-mutant glioma in children starting at 14 years of age (*41*). Given the slow growth of LGG, it is conceivable that these tumors may indeed initiate undetected in early childhood.

To assess the importance of the rs55705857 SNP, we generated mouse lines with targeted CRISPR/Cas9 mutagenesis of the orthologous murine rs55705857 locus. Importantly, genetic ablation of 66 base pairs encompassing the region orthologous to rs55705857 (thereby removing the OCT binding motif) or knocking-in the G-risk allele (together with 4bp insertion destroying the PAM site) dramatically decreased latency and increased tumor penetrance in the context of mutant *Idh1*^R132H^, *Trp53* and Atrx. While these strains do not perfectly mimic the SNP, these data clearly show that the locus is important for gliomagenesis. Together with the fine-mapping of the risk allele in human LGG and the differential affinity of the risk allele for OCT transcription factors, our data strongly suggest that rs55705857 is the causative allele.

While several other germline SNPs are associated with the development of LGG, rs55705857 confers by far the greatest risk for LGG development above and beyond combinations of the other LGG risk loci (*3-5, 42, 43*). However, the molecular basis for the rs55705857-LGG association was unknown until this study. Here, we reveal a functional link between the rs55705857 germline variants, OCT2/4-mediated regulation of MYC expression and the development of IDH-mutant LGG. Our model helps to further understand the biology of IDH-mutant gliomas and explains much of the inherited risk of developing these tumors. Importantly we have developed faithful preclinical model that can be used to assess new therapeutic avenues IDH-mutant glioma.

## Data Availability

All data produced in the present study are available upon reasonable request to the authors.

https://www.ncbi.nlm.nih.gov/geo/query/acc.cgi?acc=GSE167806

https://www.ncbi.nlm.nih.gov/geo/query/acc.cgi?acc=GSE172391

https://www.ncbi.nlm.nih.gov/geo/query/acc.cgi?acc=GSE172390

## Acknowledgements

We would like to thank all members of our laboratories as well as The Centre for Phenogenomics (TCP) for helpful discussions. We thank the staff of the Epigenomics Development Laboratory and Recharge Center (EDL) at Mayo Clinic for carrying out the epigenomic assays. The EDL is supported in part by the Mayo Clinic Center for Individualized Medicine. The authors wish to acknowledge study participants, the clinicians and research staff at the participating medical centers and the UCSF Neurosurgery Tissue Bank and Mayo NeuroOncology Tissue banks. We also thank the following colleagues at the Mayo Clinic and UCSF who facilitated subject recruitment and collecting and curating subject data: M. Bublitz, J. Buckner, T. Burns, A. Caron, C. Giannini, B. O’Neill, I. Parney, C. Praska, A. Ragunathan, J. Sarkaria, M. Berger, P. Bracci, S Chang, H. Hansen, L. McCoy, A. Molinaro, M. Prados, T. Rice, T. Tihan and J. Wiemels. We especially want to thank the many other neurosurgeons at Mayo who collected, over several years, the tissues used in this study.

## Author Contributions

C.Y. performed all mouse experiments. K.L.D. together with T.K. performed all the analysis of the human glioma samples. R.T. and J.H. helped with ChIP- and RT-PCR, M.L. helped with 4C experiments, J.P. helped with metabolomics, T.K. performed the fine mapping analyses of the 8q24 region and A.A. and T.K. performed the Mayo and TCGA GSEA analyses. P.D. and M.K. performed all the Mayo Clinic and TCGA human RNAseq studies and statistical analyses, L.W., A.A. and A.P. helped with the ATACseq, ChIPseq and RNAseq analyses, A.M. performed all bioinformatic mouse analysis, A.H., D.T. and JW helped with mouse experiments, S.K.L. and K.N.A helped with CRISPR technologies, J.F. helped with establishing primary cell lines, P.M., M.L. and H.H.H. performed IGR and motif analysis, L.Z. and A.E. performed all histology, J.W.D. supervised metabolomics, M.W. supervised 4C experiments, Daniel Lachance (D.L.) supervised the collection of the Mayo Clinic specimens and clinical data, M.W. and J.W. provided the UCSF 8q24 case and control genotyping data, L.P., D.E.D and A.V. supervised the reporter knock-in mice experiments, M.T. and P.D. provided cell lines and experimental guidance, G.Z. and L.J. performed R/S-2HG MS, Jeanette Eckel-Passow J.E.P. supervised the statistical analysis of the Mayo Clinic human data, T.M. provided the Idh1^R132H^-mutant mice, E.K. performed and analyzed the reporter knock-in mice experiments. D.S. and R.B.J. designed the experiments and coordinated the project, and together with C.Y. and K.L.D. wrote the manuscript.

## Data and materials availability

All ChIP-seq and RNA-seq for human glioma is available at NCBI Gene Expression Omnibus

## Supplementary Information

contains all Methods, Supplementary Figs. 1-9, Supplementary Fig. legends and Supplementary Tables 1&2.

