## Supplemental Information for "A non-coding single nucleotide polymorphism at 8q24 drives IDH1-mutant glioma formation"

### Materials and Methods

##### Mayo Clinic cases and controls

Mayo Clinic glioma cases have been previously described (*3-6, 42-44*). Controls were obtained from Mayo Clinic's Biobank and have been previously described (*6, 7*). Germline DNA specimens from 337 IDH mutant glioma cases and 446 controls were used for fine mapping experiments. ChIPseq and RNAseq experiments utilized tissue from surgical resections on 83 glioma cases and 9 gliosis cases performed at Mayo Clinic between 200X and 201X. Tissue sections from the frozen specimens were reviewed by a neuropathologist to confirm histology and mark tumor location. This study was approved by the Mayo Clinic Office for Human Research Protection.

| | Glioma vs Gliosis | IDH-WT | IDH-mut | | Total |
|---|---|---|---|---|---|
| Mayo RNAseq | | | codel | nonc-odel | |
| | Glioma | 26 | 29 | 26 | 83 |
| | Gliosis | 9 | | | 9 |
| ChromHMM | Glioma vs Gliosis | IDH-WT | IDH-mut | | Total |
| | | | codel | non-codel | |
| | Glioma | 9 | 26 | 22 | 57 |
| | Gliosis | 9 | | | 9 |

| | Histone mark | IDH-WT | IDH-mut | | Total |
|---|---|---|---|---|---|
| Histone ChIP | | | codel | non-codel | |
| | H3K27ac | 11 | 28 | 24 | 63 |
| | H3K4me$^1$ | 9 | 25 | 23 | 57 |

##### UCSF glioma cases

UCSF glioma cases include participants of the San Francisco Bay Area Adult Glioma Study (AGS). Details of subject recruitment for AGS have been reported previously (*3-6, 42-44*). Germline DNA specimens from 285 IDH mutant glioma cases and 222 controls were used for fine mapping experiments. This study was approved by the UCSF Committee on Human Research.

##### Haplotypes and Recombination mapping

Risk and ancestral non-risk haplotypes were constructed using phased genotyping data for SNPs from 503 EUR subjects from the 1000 Genomes project. SNPs with an alternate allele frequency <0.05 in the EUR population were excluded. Data from linked-read (10X Genomics) sequencing for 6 individuals confirmed the presence of both computed haplotypes in cases and controls (see following section for linked-read sequencing methods). Additionally, no rearrangements or novel variants were identified within the minimal association region using the 10X Genomics data.

Genotyping data from a modified Illumina OncoArray (Melin et al, 2017) was used to genotype germline DNA from 622 IDH mutant glioma cases and 668 controls (Eckel-Passow et al, 2020). Haploview (https://www.broadinstitute.org/haploview/haploview) was used to examine the haplotype structure of the region surrounding rs55705857. Guided by the Haploview results, subjects carrying the risk haplotype were identified and the genotyping data for these cases was manually examined to identify meiotic crossovers that disrupted the risk haplotype. *IDH* mutant cases and controls heterozygous for the risk haplotype that did not contain crossovers within the region were not included (n = 155 and 45, respectively). Subjects homozygous for the risk haplotype (n = 15 all of which are *IDH* mutant cases) and cases and controls not containing the risk haplotype within this region (n = 397 and 601, respectively) were also not included.

**Linked read sequencing using 10X Genomics Platform**

DNA for five subjects was extracted from 200 ul of snap frozen buffy coats using Qiagen's MagAttract HMW kit Further enrichment for long DNA fragments using the Blue Pippin system and library preparation using the 10X Genomics platform to prepare sequencing libraries was performed by GENEWIZ. The libraries were sequenced on Illumina HiSeq to ~30 fold coverage, data was processed through the 10X Genomics pipeline and the resulting data examined using the Loupe browser 2.1.1 ((https://support.10xgenomics.com/genome-exome/software/downloads/latest). Our analysis also included the sequencing data for a CEU subject, NA12878, that carries the rs55705857 G allele (Loupe file provided by 10X Genomics).

**Frozen Glioma RNA sequencing**

Frozen glioma or gliosis samples were crushed in a liquid nitrogen-chilled Cellcrusher™ tissue pulverizer (Cellcrusher, Portland, OR) and transferred to QIAzol Lysis Reagent prior to extraction with the RNeasy Lipid Tissue Mini Kit (Qiagen) per manufactures guidelines. Isolated RNA was DNase treated, ribo-depleted with the Ribo-Zero rRNA removal kit (Illumina). RNA integrity was assessed with the Agilent Tapestation. Samples were sequenced on the Illumina HiSeq in a 2x150bp configuration with a single index.

**Frozen Glioma Chromatin immunoprecipitation-sequencing (ChIP-seq)**

ChIP-seq was performed as previously described (*45*). Briefly, about 100 mg of frozen tumor or gliosis tissues were homogenized in PBS buffer. The homogenates were fixed to final 1% formaldehyde for 10 min at room temperature, quenched with 125 mM glycine for 5 min at room temperature, and followed by washing with TBS buffer. The homogenized pellets were resuspended in cell lysis buffer (10 mM Tris-HCl, pH7.5, 10 mM NaCl, 0.5% NP-40) and incubated on ice for 10 min. The lysis step was repeated. The lysates were divided into 2 tubes and washed with the MNase digestion buffer (20 mM Tris- HCl, pH7.5, 15 mM NaCl, 60 mM KCl, 1 mM $CaCl_2$). After resuspending in 250 ul of the MNase digestion buffer with proteinase inhibitor cocktails, the lysates were incubated in the presence of 1,000 Gel units of MNase (NEB, M0247S) at 37°C for 20 minutes with continuous mixing in thermal mixer (Fisher Scientific, 05-450-206). The reaction was stopped by adding 250 ul of 2X ChIP buffer (100 mM Tris-HCl, pH 8.1, 20 mM EDTA, 200 mM NaCl, 2% Triton X-100, 0.2% sodium deoxycholate). The chromatin was further sonicated using a Bioruptor sonicator (Diagenode, Denville, NJ) by 30 cycles (30s on and 30s off) and soluble chromatin was isolated by centrifugation (21,130 x g, 10 min). Chromatin input was incubated with the antibodies against $H3K4me^1$ (CST, Cat# 5326, Lot 1), $H3K4me^3$ (EDL, Lot 1), H3K27ac (CST, Cat# 8173, Lot 1), or $H3K36me^3$ (Active motif, Cat# 61101, Lot 32412003) overnight at 4 °C and precipitated using Protein G-magnetic beads (GE Healthcare Life Sciences, Pittsburgh, PA). The beads were extensively washed with ChIP buffer, high salt buffer, $LiCl_2$ buffer, and TE buffer. Bound chromatin was eluted and reverse-crosslinked at 65°C overnight. DNAs were purified using Min-Elute PCR purification kit (Qiagen) after treatment of RNase A and proteinase K. The enrichment was analyzed by targeted real-time PCR in positive and negative genomic loci. The size of input chromatin was determined using an Advanced Analytical Technologies (Ankeny, IA) Fragment Analyzer™ Automated CE System. The libraries were prepared from 1 - 10 ng of ChIP and input DNAs with the ThruPLEX DNA-seq Kit (Rubicon Genomics, Ann Arbor, MI) and were sequenced (paired-end; 51 bases/read) on the Illumina HiSeq 4000 platform in the Mayo Clinic Medical Genomics Core.

**RNAseq preprocessing and statistical analysis**

MAP-RSeq workflow was used to process the RNA sequencing data (*46*). STAR (*47*) was used to align reads to the reference human genome, build hg38. Gene and exon expression quantification was performed using the Subread package (*48*) to obtain both raw and normalized (RPKM – Reads Per Kilobase per Million mapped) reads. Comprehensive quality control analyses from the RSeQC (*49*) package was used to assess the quality of the sequenced libraries. Differential expression of the gene counts by rs55705857 and tumor/tumor comparisons was performed using the R package edgeR (*50, 51*). Unsupervised hierarchical clustering was performed using the R package heatmap3 (*52*). Both human and mouse samples were included in the clustering. The clustering was limited to transcripts included in both the human and mouse sequencing. Differential expression was performed between mouse and human samples using edgeR and transcripts that were significantly different ($p<0.01$) were removed. The median absolute deviation was then calculated on the RPKM value of the remaining transcripts and the log of the top 1000 most variable transcripts were used for the clustering. One minus the Pearson correlation coefficient was used as the distance metric for the clustering. TCGA LGG RNAseq data was downloaded from the *recount2* online resource (*53*). Differential expression of the TCGA data was performed by rs55705857 and tumor/tumor using edgeR and results were compared to the analysis of the Mayo data to determine which transcripts validated.

**Human GSEA**
Pre-ranked lists of genes were generated from the RNAseq edgeR analysis (see RNAseq preprocessing and statistical analysis) by combining the sign from the log fold change and the F statistic to produce a signed F statistic. Gene lists were filtered to remove genes with median counts <16. GSEAv4.1.0 (https://www.gsea-msigdb.org/gsea/index.jsp) was used to perform pre-ranked GSEA analysis using the protocol published by Reimand et al. (*54*) to examine the Hallmark 50 gene sets.

**TCGA ATAC-seq Analysis**
ATAC-seq data (bigwig-files with per base coverage across GRCh38 reference genome) for LGG (13 samples), GBM (9 samples) and SKCM (13 samples) was taken from TCGA (Corces *et al.*, 2018, https://gdc.cancer.gov/about-data/publications/ATACseq-AWG). We stratified the LGG and GBM samples based on their IDH mutation (i.e. R132) status and subdivided the IDH mutation samples further based on presence of 1p-19q co-deletion. The molecular and copy number information were taken from published data (*55*). In LGG cancer type, 1 sample has IDH-WT, 7 samples have IDHmut non-codeletion and 5 samples has IDH-mut codel. For GBM cancer type, 8 samples have IDH-WT and 1 sample has IDH-mut non-codel.

Summary of the utilized ATAC-seq data from TCGA.

| Cancer Type | IDH-WT | IDH-mut | | Total |
|---|---|---|---|---|
| | | co-del | non-codel | |
| LGG | 1 | 5 | 7 | 13 |
| GBM | 8 | 0 | 1 | 9 |
| SKCM | 13 | | | 13 |

Differential binding analysis was conducted to compare the peaks between groups. Samples were grouped based on their IDH status from LGG and GBM cancer type. Comparison is done between IDH-WT vs IDH-Mut. DiffBind (*56*) program was used for this analysis and within this DESEQ2 (*57*) method is used to compare the peaks for finding the differentially bound sites between compared groups.

**Human Chromatin immunoprecipitation sequencing (ChIP-seq) data analysis (histone mark peak calling, diffbind, ChromHMM)**
ChIP-seq data were analyzed using the Mayo Clinic HiChIP workflow (*58*). Specifically, 51mer, paired-end reads were first analyzed for quality using FASTQC (https://www.bioinformatics.babraham.ac.uk/projects/fastqc/) and then aligned to the human reference genome (GRCh38/hg38) using BWA (version 0.7.17-r1194). The resulting BAM files were sorted and indexed with Samtools (version 0.1.12a) (*59*). PCR and optical duplicated reads were marked and removed using Picard tools (version 1.67) (https://broadinstitute.github.io/picard/). Narrow peaks (H3K4me$^1$, H3K4me$^3$, H3K27ac) were called by MACS2 (version 2.0.10.09132012) with q-value cutoff 0.01 (*60*). Broad peaks (H3K36me3) were called by SICER (version 1.1) with FDR cutoff 0.01 (*61*). CEAS and

GREAT were used to visualized and annotate (*62, 63*). FRiP (fraction of reads in peak) score was calculated using deeptools (*64*). BigWig files were generated for visualization with the UCSC genome browser or IGV.

ChromHMM analysis was performed on 4 histone marks (including H3K4me$^{1}$, H3K4me$^{3}$, H3K27ac, and H3K36me$^{3}$) of 77 patients (*65*). Duplicate reads in the BAM files were collapsed. Binarization was performed on BAM files of ChIP samples and the corresponding input controls were used as additional features to adjust the local binarization threshold. We set the '-f' to 3, and used the defaults for other options. During model learning, we set 'numstates' to 8. BigBed files were generated to visualize the chromatin states of each patient.

A total of 10 differential binding analyses were performed for each of the four histone marks (https://bioconductor.org/packages/release/bioc/html/DiffBind.html). Differential peaks were called based upon a p-value less than 1e-5 with a log2 fold change with absolute value greater than 1. Differential peaks were annotated and assigned to the putative target genes by the HOMER ( v4.10) (*66*).

**Animals**

Equal numbers of male and female animals were used throughout the study without any bias, with the exception of transgenic mouse embryos, which included male and female embryos, but were not examined/scored separately by sex. Animal husbandry, ethical handling of mice and all animal work were carried out according to guidelines approved by Canadian Council on Animal Care and under protocols approved by the Toronto Centre for Phenogenomics Animal Care Committee (18-0272H). All mouse work performed at Lawrence Berkeley National Laboratory was reviewed and approved by the Lawrence Berkeley National Laboratory Animal Welfare and Research Committee. The animals used in this study were LSL-Idh1$^{R132H}$, kindly provided by Dr. Tak Mak and described previously (*19, 67*)), R26-LSL-Cas9-GFP (JAX #026175), Trp53$^{LSL\text{-}R270H}$ (#129S-Trp53$^{tm3Tyj/J}$, JAX #008651), Trp53$^{fl/fl}$ (Trp53$^{tm1Brn}$, JAX #008462), Ptenfl/fl (Pten$^{tm1Hwu/J}$, JAX #006440) obtained all from Jackson laboratories. Atrx$^{fl/fl}$ mice (Atrx$^{tm1Rjg}$, MGI ID: 3528480) were kindly provided by Dr. David Picketts. Notch1$^{fl/fl}$ mice were described previously (*68*). The rs557$^{A\text{->}G_4bp}$ and rs557$^{66bp}$ mutant lines were generated in the Toronto Centre for Phenogenomics. Genotyping was performed by PCR using genomic DNA prepared from mouse ear punches.

**Enhancer-reporter assays in mouse embryos**

A human hs1709 enhancer (chr8:129,631,847-129,635,071; hg38; 3225 bp) containing the reference or the variant allele was PCR-amplified from DNA from the 1000 Genomes Project and cloned into the PCR4-Shh::lacZ-H11 plasmid (Addgene plasmids #139098) upstream of a minimal *Shh* promoter and *lacZ* reporter gene (Kvon et al., 2020). The HG00310 DNA sample for PCR amplification of the variant allele was obtained from the NIGMS Human Genetic Cell Repository at the Coriell Institute for Medical Research. We used a mouse enhancer-reporter assay that relies on site-specific integration of a transgene into the mouse genome (Kvon et al., 2020). Transgenic assays were performed in the FVB mouse strain. Embryos with integrated enhancer-reporter transgene were collected and LacZ-stained at two time points, E11.5 and E14.5. Embryos were excluded from further analysis if they did not carry the reporter transgene integrated at the H11 locus. All embryo images for the reference and variant alleles of hs1709 enhancer were randomized, the labels removed, and annotation was performed by at least four independent reviewers blinded to the genotype. For each embryo, reviewers scored whether the enhancer activity pattern in each tissue was 1) robust, 2) weak, or 3) absent. Final annotations were determined by the staining type with the most reviewer votes.

**Generation of mice with mutations in the homologous rs55705857 loci**

The human genomic sequence surrounding rs55705857 was aligned with the syntenic regions of the mouse and human genome to identify the target nucleotide. The target A nucleotide is conserved between mouse and human and adjacent sequences are only slightly divergent:

```
Human 8:129633407    ATGAGGAATTCCAAGGACTTG-CTCAGAAGCCGTTCTGCARATGCATTTCTTGAACAAAG
```

```
Mouse 15:63675539   ATGAGGAATTCCAAGGACTTAACTCAGAAGCCGCTCTGCAAATACATGGCTTGAACAAAG
                    ********************. *********** ****** **.*** ***********
```

The desired allele is a A>G mutation at Chr15:63675579. Sequence analysis of the ~70 bp surrounding this site in genomic DNA revealed gRNAs sequence with predicted endonuclease sites at or near Chr15:63675579. These gRNA sequences were scored for specificity (*69*). Using this prediction algorithm, gRNA_ag appears to have a suitable target endonuclease site – 2 bp from Chr15:63675579 – with good specificity. All putative off-target sites have at least 3 mismatches:

| gRNA | Sequence | Location (Strand) | Specificity Score | No. off-target sites | |
|---|---|---|---|---|---|
| | | | | Total | In genes |
| gRNA_ag | GAAGCCGCTCTGCAAATACA | Chr15: 63675565 (+1) | 72 | 172 | 17 |

The specificity score of each gRNA is predicted to reflect the faithfulness of on-target activity and is calculated as 100% minus a weighted sum of off-target site scores in the target genome (Mus musculus) (http://crispr.mit.edu/about). The off-target site scores are calculated using an algorithm that takes into account the experimentally determined effect of gRNA:target DNA mismatches. Using Cas9 to create mutations in mouse zygotes demonstrated an undetectable off-target rate for gRNAs with ≥3 mismatches at all putative off-target sites (0 off-target mutations from among the top 8-10 scoring off-target sites for 64 different gRNAs screened in 207 N1 mice – a total of 3,060 loci were screened). We used a repair template will be a single-strand oligonucleotide encoding the A>G change (red boldface):
TTGGCACGGTCCTAATGACTTAAAGAAAATGCCGGAAACCAGCCCCGCACTCTTTTCTGAAAGGAACATTGGCCCTTTGTTCAA
GCCATGTAT**C**TGCAGAGCGGCTTCTGAGTTAAGTCCTTGGAAT
This mutation introduces a single base-pair change in the seed region of the gRNA, but does not disrupt the protospacer adjacent motif (PAM). The oligonucleotide is antisense to the PAM-containing strand and in an asymmetric oligonucleotide design to improve repair frequency (*70*).

The CRISPR/Cas9 sgRNA RNP was inject WT B6J zygotes and targeted founder mice were identified using sanger sequencing of a 344bp PCR amplicon encompassing the homologous rs55705857 locus using the rev. primer (fwd. primer: GAAAGCCCTTGCTTCTTCAATAC and rev. primer: CTGTCTGTAGCCACCATCAATAA). Two founder alleles were identified (Figure S6C) and backcrossed three times to B6J mice before intercrossing to the experimental $Idh1^{LSL\text{-R132H/+}}$;$Atrx^{fl/fl}$;$Trp53^{fl/fl}$;R26-Cas9 mice.

**Lentiviral constructs**

All the sgRNAs targeting gene of interest were cloned into pLKO-Cre stuffer v4 plasmid (Addgene #158032) (*71, 72*) by using BsmBI restriction sites. sgRNAs targeting genes found mutated in LGG and non-targeting sgRNAs were ordered as oligos from IDT (Table S3 sgAtrx #1: CTTGTCATGAACCAAAGCAC, sgAtrx #2: GTGCTGAATGAAGACAAAGA) and cloned into pLKO-Cre stuffer v4 using BsmBI restriction sites.

**Lentivirus production and transduction**

Large-scale production and concentration of lentivirus were performed as previously described (*71, 72*). 293T cells (Invitrogen R700-07) were seeded on a poly-L-lysine coated 15 cm plates and transfected for 8 hours using PEI (polyethyleneimine) method in a non-serum media with lentiviral construct of interest along with lentiviral packaging plasmids psPAX2 and pPMD2.G (Addgene #12259 and 12260). Post transfection, media was added to the plates supplemented with 10% Fetal bovine serum and 1% Pencillin-Streptomycin antibiotic solution (w/v). 48 hours after transfection, viral supernant was filtered through a Stericup-HV PVDF 0.45-µm filter, and then concentrated ~2,000-fold by ultracentrifugation in an MLS-50 rotor (Beckman Coulter). Viral titers were determined by FACS based quantification of infected R26-LSL-tdTomato MEFs.

**Adenoviral constructs**

Adenovirus (Ad-Cre and Ad-GFP) was ordered from University of Iowa Viral Vector Core Facility.

**Intercranial injection and lentiviral transduction**

As previously described (*73, 74*), virus was mixed with 0.05% Fast Green (F7252-5G) and loaded into a syringe (Hamilton 7659-01) with 33-gauge needle (Hamilton 7803-05). P0 pups were anesthetized on parafilm covered ice and their head secured with a custom 3D-printed mold. A stereotactic manipulator was used to position the needle to 0.3 mm above the Bregma towards the Lambda Suture and 0.1 mm lateral of Sagittal Suture into the right ventricle. The needle punctured 3 mm into the skull, and retracted 1mm for a final depth of 2 mm. 1 μl of virus was administered and allowed 1 minute to diffuse before retraction of the needle. Post-injection, the neonates were warmed by cupping them in hands until conscious.

**2HG-Sample quantification**

One 10 cm plate of confluent $Idh1^{LSL\text{-}R132H/+}$ primary NSC or 500mg brain tissue of $Idh1^{LSL\text{-}R132H/+}$ mice transduced with either Ad-GFP or Ad-Cre-GFP was harvested, pelleted and snap frozen at -80 °C. Samples were analyzed by LC/MS/MS at the Analytical Facility for Bioactive Molecules (The Hospital for Sick Children, Toronto, Canada). Sample extraction was prepared based on published method (Dang et al., 2009) with slight modifications. Briefly, 200 µL of ultrapure water were added to cell pellets. Samples were sonicated for 1 minute on ice (550 Sonic Dismembrator, Fisher Scientific). 20 µL were transferred into a set of 1.5 mL Eppendorf tubes and volume was adjusted to 200 µL with ultrapure water. 800 µL methanol were added, samples were mixed by vortex for 5 minutes and kept on ice for 15 minutes. After centrifugation (20,000 g, 15 minutes at 4°C) to remove precipitated proteins, supernatants were dried under a stream of nitrogen. Residues were reconstituted in 200 µL methanol/water (1/1) and after centrifugation (20,000 g, 15 minutes at 4 °C), clear supernatants were transferred into a set of 200 µL plastic inserts for LC-MS-MS analysis.

2-HG is measured by liquid chromatography-tandem mass spectrometry using a QTRAP 5500 triple-quadruple mass spectrometer (Sciex: Framingham, Massachusetts, USA) in negative electrospray ionization mode by MRM data acquisition with an Agilent 1200 HPLC (Agilent Technologies: Santa Clara, California, USA ). Chromatography is performed by automated injection of 5 µL on a Synergi hydro RP column, 150 x 2 mm, 4.0 µm particle size) (Phenomenex, Torrance, CA). The HPLC flow is maintained at 300 µL/minute with a gradient consisting of: A= 10 mM ammonium acetate, pH 3.2 and B = methanol. Total run time was 10 minutes.

**R/S-2HG quantification**

Quantification was done on Analyst 1.6.1 software (ABSciex : Framingham, Massachusetts, USA) by plotting the sample peak area ratios (Analyte peak area/Internal Standard peak area) of 2-HG against a standard curve generated from various concentrations of 2-HG from 1 ng to 750 ng, spiked with the same amount of 2-HG-d4 used for the samples and extracted in the same conditions

**Metabolomic Profiling by LC/MS**

Mouse Neural stem cells 500,000 were seeded in 6-well plates and cultured for 48 hours. Media was aspirated, cells washed with warmed PBS (Wisent, 311-010-CL) and then snap frozen in liquid nitrogen. Metabolites were extracted by adding 1mL of extraction solvent (40% acetonitrile, 40% methanol and 20% water), scraped and transferred to a 1.5mL tube. Samples were shaken at 4°C at 1400 rpm for 1 hour. Samples were spun down and supernatant was placed into a new tube where it was evaporated in a CentreVap concentrator at 40°C. Samples were stored at 80°C for later LC-MS/MS analysis.

Dry extracts were reconstituted in 100ml of water containing internal standards (500mg/mL and 300mg/mL of D7-glucose and 13C9 15N-tyrosine). The metabolites were separated at room temperature through a guard column (Inertsil ODS-3, 4 mm internal diameter X 10 mm length, 3mM particle size) and analytical column (Inertsil ODS-3, 4.6 mm internal diameter, 150 mm length, and 3-mM particle size) for both polarity modes. Eluted metabolites were analyzed at the optimum mass spectrometric conditions as published (*75*). Peaks were manually confirmed in Multiquant (*75*) and area ratio compared to IS was normalized to final cell counts and uploaded for statistical analysis in MetaboAnalyst (*76*).

**Tide analysis**

LSL-Cas9-GFP MEFs were cultured and infected with lentivirus carrying Cre and corresponding sgRNAs. Cells were live sorted for GFP expression and expanded further to extract genomic DNA using DNeasy Blood & Tissue Kit (Qiagen). Genomic DNA from tumors from the mice injected with single sgRNAs were also isolated using the same kit. PCR was performed flanking the regions of sgRNA on genomic DNA from both WT MEFs and cells infected with respective virus or tumors and sent for Sanger sequencing. Sequenced chromatograms were uploaded to https://www.deskgen.com/landing/tide.html and genome editing efficiency was estimated.

**Western Blot analysis**

Protein extract from cell lysates were harvested by adding RIPA lysis buffer directly confluent 6-well cultures of either RIP or mouse NSCs at on ice for 15 minutes. Cell lysates were then transferred to an Eppendorf and spun at 16,000xg for 15 minutes and supernatant was transferred a new Eppendorf tube. Samples were boiled in loading dye with 1/50 volume 2-Mercaptoethanol for 5 minutes and loaded into a Bolt BisTris Plus 4-12% gradient gel (Life Technologies, NW04127BOX) with Bolt MOPS SDS Running Buffer (Life Technologies, B0001). Protein gels were run for 40 minutes at 200V, and subsequently transferred to a PVDF membrane (Millipore, IPFL00010) in Tris Glycine buffer (Wisent, 880-560-LL) for 2 hours at 0.3 amps. Western blots were blocked in 5% milk for 1 hour and placed in primary antibody diluted in 5% milk over night at 4°C. Primary antibodies used were 1:1000 diluted Idh1$^{R132H}$ (generous donation of MilliporeSigma, 456R-31), 1:10000 diluted Gapdh (Biolegend, 607904), 1:1000 diluted Oct2 (Life Span Biosciences, LS-C124305-100) and 1:1000 diluted Oct4 (Abcam, ab181557). All Blots were washed with PBS and then placed in Goat Anti-Rabbit IgG HRO (Bio Rad, 1706515) diluted 1:10000 in 5% milk for 1 hour. Blots were washed with PBS before adding 500ul of Clarity Western ECL Substrate (Bio Rad, 1705061) and imaging on a Bio Rad ChemiDoc XRS+.

**Mouse brain tissue histology**

Brains of mice were dissected and submerged in 10% buffered formalin for 2-3h, then cut midline sagittal, and continued fixation for at least 48h, after which they were transferred to 70% ethanol and subsequently processed and embedded in paraffin following standard procedures. Brains were oriented in a “butterfly” pattern and sections cut with two sections per slide for staining. Prior to staining, slides were heated at 60oC for 15min, then dewaxed and rehydrated. Slides for immunohistochemistry were treated with 3% hydrogen peroxide in PBS for 15min to kill endogenous peroxidases. (For the x-Nestin antibody to work required killing of endogenous peroxidases with 10% H2O2 in 70% MeOH.) Slides were then washed in PBS followed by microwave antigen retrieval using NaCit (pH6) or TRIS-EDTA (TE, pH9). Only x-GFAP antibody did not require AgRet. The following markers and conditions were used to evaluate our thesis: IDH1$^{R132H}$ (generous donation of MilliporeSigma, 456R-31), TE/1:400; PDGFRa (R&D AF1062), TE/1:100; Ki67 (Abcam ab15580), NaCit /1:1000; GFAP (Dako Z0334) no AgRet/1:200; Olig2 (Millipore AB9610), TE/1:500; Nestin (Pharmingen 556309), TE/1:150; Cleaved Caspase 3 (ASP175 – Cell Signalling Technologies 9661), NaCit/1:750; and p53 (CM5, Vector Labs VP-P956), NaCit/1:1000. Secondary antibodies (Vector Labs) were diluted in PBS and 0.2% Triton X-100 applied at 1:500, (x-rabbit, BA-1000; x-goat, BA-5000; x-mouse, BA-9200). An ABC kit (Vector Labs, PK-4100) and DAB Reagent (Vector Labs, SK-4100) were used according to the manufacturer’s instructions. Primary antibodies were blocked and diluted in a Histoblock solution with 5% serum of the host of the secondary antibody and 0.2% Triton X-100. Primary antibodies were incubated at room temperature for 35-45minutes, secondary antibodies for 35min, and ABC for 25minutes. The DAB reaction was allowed to proceed for 2-4minutes. Slides were counterstained for 8 min in Harris Hematoxylin, washed in water, dehydrated, and mounted in a xylene based mounting medium. Stained sections were digitized at 40x using a Hamamatsu Nanozoomer Scanner (2.0-HT).

**RNAseq of mouse tumor tissue and GSEA**

RNA quality was assessed using an Agilent 2100 Bioanalyzer, with all samples passing the quality threshold of RNA integrity number (RIN) score of >8. The library was prepared using an Illumina TrueSeq mRNA sample preparation kit at the LTRI sequencing Facility, and complementary DNA was sequenced on an Illumina Nextseq platform. Sequencing reads were aligned to mouse genome (mm10) using Hisat2 version 2.1.0 and counts were obtained using featureCounts (Subread package version 1.6.3). Differential expression was performed using DESeq2 release 3.8. Gene set enrichment analysis was performed using GSEA version 3.0. Samples were processed using the Hallmark gene sets gathered from https://www.gsea-msigdb.org/gsea/msigdb/collections.jsp#H.

**Derivation and culturing of mouse tumor cells**

Tumour tissue was harvested and washed in HBSS (Wisent, 311-512-CL) before transferred into 10cm plate containing 10ml TrypLE Express (Life Technologies, 12604021) and incubated for 10 minutes at 37°C. Tumour tissue was then mechanically dissociated with a pipette and transferred to a 50ml conical tube with 40ml HBSS. Samples were centrifuged at 300xg for 5 minutes, supernatant was transferred to a new 50ml conical tube and spun at 300xg for 5 minutes. Both cell pellets were pooled and washed with 40ml of HBSS. The pellet was resuspended in Tumour Stem Medium Base Formulation (Wisent, 305-485-CL) and seeded into 6-well plates +/- 2uM AG120.

Mouse tumour cells were cultured in at 37°C, 5% $O_2$ and in Tumour stem medium base formulation supplemented with 1X concentration W21 (Wisent, 003-017-XL), 1X concentration N2 (Wisent, 305-016-IL), 20ng/mL H-EFG (Wisent, 511-110-EU), 20ng/mL H-FGF basic (Wisent, 511-126-EU), 10ng/mL H-PDGF-AA (Shenandoah, 100-16-100UG), 10ng/mL H-PDGF-BB (Shenandoah, 100-18-100ug) and 2ug/mL heparin solution (Stem cell technologies, 07980). Cells were passaged using TrypLE Express.

**ChIP-PCR / RT-PCR**

Chromatin Immunoprecipitation was performed with the SimpleChIP Enzymatic Chromatin IP Kit (#9003, Cell Signaling Technology) according to the manufacturer's protocol with minor modifications. Cells cross-linking for ChIP: Confluent 10 cm plates of RIP cells and RIP cells treated with OCT4 were used for each experiment. Cells were cross-linked while in media with 270 uL of 37% formaldehyde. Following a 10 min incubation at room temperature, 1 mL of 10X glycine was added to each plate. After 5 minutes of incubation at room temperature, cells were washed twice with ice cold PBS and scraped into cold PBS buffer containing protease inhibitor cocktail. Harvested cells were frozen in -80°C until all samples have been processed before starting the next step. Chromatin extraction: Cross-linked cells were thawed on ice and resuspended in 1 mL cold Buffer A, mixed well, and centrifuged at 2000x g for 5 min at 4°C. The pellet was then mixed in 1 mL cold Buffer B, incubated on ice for 10 min, then centrifuged at 2000x g for 5 min at 4°C. After resuspension in Buffer B, chromatin was digested with 0.5 uL MNase for 20 min at 37°C, using a Thermomixer at 850 rpm. Finally, the reaction was stopped by the addition of 10 uL 0.5 M EDTA, and the tubes were then placed on ice for 2 minutes. The cells were then pelleted and resuspended in 400 uL cold ChIP buffer, and sonicated on ice using a 3.2 mm microtip with QSonica Q700 (25% amplitude, 10 seconds on, 15 seconds off) for a total of 3 cycles to release the chromatin. All the lysates were centrifuged at 9400x g for 10 min at 4°C. The supernatant was collected and total chromatin quantified as per kit instructions before the start of each immunoprecipitation (IP). Immunoprecipitation: Total digested chromatin was diluted accordingly to a total volume of 500 uL, with 2 ug of chromatin per IP, in cold ChIP Buffer. Each ChIP sample was precleared by incubating with 30 uL of Protein G magnetic beads, at 4°C with rotation for 1 hour. Samples were then incubated with the appropriate antibody (2 ug per IP) at 4°C overnight with rotation. We prepared MNase ChIP-seq data using the following antibodies: H3K4me$^1$ (ab8895, Abcam, lot: GR3312607-1), H3K27ac (ab4729, Abcam, lot: GR3187598-1), OCT4 (ab181557, Abcam, lot: GR3236296-3), SOX2 (AF2018-SP, R&D Systems, lot: KOY0419061) and OCT2 (C124305, LifeSpan BioSciences, lot: 167932). OCT4 antibody (sc9081, Santa Cruz, Lot: B0113) was used for human cells. All buffers and solutions, including antibodies against total histone H3 protein (H3KPan) and normal rabbit IgG, were provided by Cell Signaling Technology (#9003 Simple ChIP kit). On the following day, 30 uL of Protein G magnetic beads were added to each sample and then further incubated at 4°C with rotation for 2 hours. After incubation, samples were placed on a magnetic rack and washed three times with 1 mL Low Salt Wash Buffer for 5 min at 4°C with rotation, then washed once with High Salt Wash Buffer for 10 min at 4°C with rotation. The remaining beads were resuspended in 150 uL Elution Buffer and incubated in a Thermomixer at 1200rpm for 30 min at 65°C. The eluted fractions were then treated with 6 uL 5M NaCl and 2 uL Proteinase K, and incubated overnight at 65°C to reverse the cross-linker. Finally, samples were cleaned up according to manufacturing instructions and purified samples were used for further analysis. Quantification by RT-PCR and PCR: A set of primers were specifically designed to flank the cassette insertion region in the targeting homologous mouse rs557055857 locus (Fwd: 5'-ACCGGGAGCTTACAAAGACA-3', Rev: 5'-ACATTGGCCCTTTGTTCAAG-3'). The same primer set was used for both RT-PCR and PCR quantification. The WT product size is 117bp, whereas the rs557 knock out product size is 51bp. For human LGG lines, we used (Fwd: 5'-TTCTTCAATGCCAGGAGCTT-3', Rev: 5'-AAAGAAAAATGCCGGAAACC-3'). A mastermix was created as follows: 3 uL nuclease-free water, 5 uL 2X PowerUp SYBR Green Master Mix (A25742, Applied Biosystems, lot: 00828561), 1 uL of 5 uM rs557 primer mix and 1 uL of purified DNA. For PCR, samples went through the PCR reaction program according to manufacturing instructions with a total of 32 cycles and products were ran on a 2.5% gel. For RT-PCR, samples went through the RT-PCR program as per kit protocol in a CFX384 machine (Biorad). Each reaction was performed in technical triplicates. Enrichment over IgG was calculated by using threshold cycles for each sample normalized to the IgG control after setting the input value at 2%.

**CRISPR interference**
IPO cells were transduced with low-titter virus expressing dCas9 protein and blasticidin resistance (Addgene: Plasmid #89567) and spun in a 37°C centrifuge at 1100xg for 1 hour. Cells were recovered for 48 hours post transduction before undergoing blasticidin selection for one week. After blasticidin selection (Wisent, 450-190-XL), IPO dCas9 cells were transduced with pLKO puromycin resistance sgrs557 or sgNTC constructs and underwent puromycin selection (Wisent, 450-162-XL) for one week. Cells were then cultured without selection and harvested for RT-PCR.

**OCT4 (Pou5f1) overexpression cell lines**
Mouse RIP LGG cells were transduced with pLEX306-Oct4V5-Cre and Oct4 expression was monitored through Western blot Analysis 48 hours post transduction. After 24hr, cells were fixed directly on the cell culture plate with 1% formaldehyde processed for ChIP.

**Circular Chromosome Conformation Capture -Seq**
Neural stem cells were cultured in 10cm plates and crossed linked with 1% PFA for 10 minutes and subsequently quenched with 2.5 M glycine and washed with PBS. 4C-seq was performed using a modified version of the 4C-seq protocol as previously described (*77*). Briefly, cell pellets were incubated on ice for 15 minutes in lysis buffer (10mM Tris-HCl pH 8, 10mM NaCl, 0.2% Nonidet P-40 and 1x protease inhibitor). Pellets were then centrifuged at 2500G at 4°C, the supernatant discarded, resuspended in 50 ul of 0.5% SDS and incubated at 62°C for 10 minutes. 170uL of 1.47% triton was added and samples were incubated at 37°C for 15 minutes. The first digestion was performed using 200U DpnII (NEB R0543L) overnight at 37°C and 700rpm. Samples were heat inactivated at 65°C and cooled to room temperature. In-situ ligation was performed at RT for 15minutes on a rotator using high concentration T4 DNA Ligase (NEB, M0202M), pelleted and resuspended in 550 uL 10mM Tris buffer pH 7.5. Chromatin was reverse-crosslinked by addition of 50ul 20mg/ml proteinase K and 57ul of 10%SDS at 55°C for 30 minutes. 67uL of 5M NaCl was added to the samples and incubated for 1 hour at 68°C. DNA was purified using the 0.8X

AmpureXP clean up (Beckman Coulter, A63880). The second digestion was performed with addition of 20U NlaIII (NEB R0125L) for 2 hours at 37°C at 700 rpm and was subsequently heat inactivated at 65°C. The second ligation was performed at 16°C overnight under dilute conditions in a total volume of 5mL per sample. DNA was then purified by adding 0.1X volume of sodium acetate, 0.7 volumes of isopropanol, 2uL of 5ug/uL Linear-polyacrylamide (Thermoscientific, AM9520) and 2uL glycoblue (Fisher Scientific, AM9515) overnight at -80°C. DNA was washed with 70% ethanol, air dried, resuspended in 200uL 10mM Tris buffer pH 7.5 and purified using 1.8X AmpureXP clean up protocol.

Primers targeting Myc were designed using the 4C primer designer (https://mnlab.uchicago.edu/4Cpd/). Viewpoint-specific primers (bold) were appended with Illumina-compatible adapter sequences (black).

Myc-upstream-reading: TCCCTACACGACGCTCTTCCGATCT**TAGACCTCATCTGCGGTTGATC**

Myc-upstream-nonreading:
GTGACTGGAGTTCAGACGTGTGCTCTTCCGATC**AAAATCAAGGCGCTAGACGC**

4C PCR was performed as previously described (*77*) using the Expand Long Template PCR System (Sigma, 11681834001) and purified using Zymogen DNA Clean & Concentrator (Zymogen, D4003). NEBNEXT adaptors and barcodes were added using PCR amplification and the samples submitted for Illumina sequencing. 4C-seq libraries were obtained in fastq format and filtered for reads containing 4C primer sequences for Myc with an edit distance of <=2. Analysis and visualization of the resultant filtered reads was performed using 4Cseqpipe (*77*). Only R1 reads were used.

For domainograms, the black trendline shows the median contact frequency in 2-kb windows tiled across 1-kb increments normalized to the maximum median value at 2-kb resolution; shaded area indicates the 20th to 80th percentiles. The heat map color-scale shows median contact frequency in the windows of increasing size from 2 to 50 kb tiled across 1-kb increments, normalized relative to the maximum median value at 12-kb resolution.

**Statistics and reproducibility**

All quantitative data are expressed as the mean ± SD. Differences between groups were calculated by two-tailed Student's t-test or one-way analysis of variance using Prism 7 (GraphPad software). $P < 0.05$ denotes significance.

**Fig. S1. rs55705857 is located in a brain-specific enhancer at 8q24**

(**A**) Roadmap and ENCODE data showing the overlap between rs55705857 (indicated by the red line) and enhancer features in 64 normal tissues and normal blood, including the brain.

(**B**) Genomic locus surrounding rs55705857 (chr8:129,608,396-129,658,495; hg38). ATACseq data from the TCGA LGG and GBM, stratified by *IDH* mutation status, are displayed, each line represents an individual tumor (dark blue tracings). The fold change between the ATACseq signal for *IDH*-mutant vs. *IDH*-WT is displayed just above the tracings (blue, 0.001>p>0.002). ChIPseq signals for H3K27ac (dark green) and H3K4me$^{1}$ (orange) are shown for the Mayo cohort of tumors, stratified into *IDH*-WT, *IDH*-mutant non-codel and *IDH*-mutant codel groups. DiffBind shows the fold change and p-value between the *IDH*-mutant vs. *IDH*-WT for both H3K27c and H3K4me$^{1}$ (blue, 0.001>P>0.002; pink, 0.002>P>0.0001; red, P<0.0001). Red line indicates position of rs55705857.

**Fig. S2. rs55705857 risk allele G enhances an LGG-transcriptional profile**
(**A**) *IDH1* mutation and ATAC-seq accessibility at rs55705857 SNP (chr8:129,633,446; hg38) and at genomic locus surrounding rs55705857 (chr8:129,630,000-129,640,000) in 377 TCGA samples across 23 cancer types using UCSC Xena. (**B**) Comparison of GSEA results using 50 Hallmark gene sets in *IDH*-mutant non-codel tumors vs. gliosis and rs55705857 A vs. G allele in *IDH*-mutant non-codel tumors.

**Fig. S3. IDH1$^{R132H}$ metabolically rewires NSC**

(**A**) Schematic of IDH1$^{R132H}$ induction in mouse NSC.

(B) Western blot of IDH1$^{R132H}$ expression in uninfected or Ad-GFP or Ad-Cre-GFP transduced LSL-*Idh1*$^{R132H}$ mouse NSC using an IDH1$^{R132H}$ specific antibody.

(**C**) Ratio of R- to S-2HG in LSL-*Idh1*$^{R132H}$ mouse NSC transduced with Ad-GFP or Ad-Cre-GFP and treated with or without AG120 measured by mass spectrometry. Statistical comparison was performed using two-tailed t-test.

(**D**) Heatmap of 79 metabolites in syngeneic *Idh1*$^{+/+}$ and *Idh1*$^{R132H/+}$ NSC cultured in 4 different glutamine concentrations measured in sextuplicate by mass spectrometry.

(**E**) Normalized concentrations of isocitrate and the α-ketoglutarate in syngeneic *Idh1*$^{+/+}$ and *Idh1*$^{R132H/+}$ NSC cultured in 4 different glutamine concentrations measured in sextuplicate.

(**F**) Bar chart of enrichment analysis of metabolic pathways altered in syngeneic *Idh1*$^{R132H/+}$ compared to syngenic *Idh1*$^{+/+}$ NSC cultured in 4 different glutamine concentrations. Scale bar at the top indicate p-value.

**Fig. S4. *In vivo* Transduction of the Mouse Brain**
(**A**) Lollipop plot depicting *TP53* mutations found in human LGG (data from the TCGA database).
(**B**) Schematic of the LSL-Cas9-GFP mouse cassette (Top). Schematic of the viral Cre injection targeting NSC of the SVZ of LSL-Cas9-GFP reporter mice, which leads to linage tracing of neural stem/progenitors and their progeny in the rostral migratory stream (RMS) and olfactory bulb (OB). Brightfield and fluorescent images of LSL-Cas9-GFP mouse brains at postnatal day 21 (P21) transduced with lentiviral Cre at P1.
(**C**) Representative immunofluorescent and bioluminescence images of LSL-Cas9-GFP brains at postnatal day 7 (P7) transduced with lentivirus (LV), adenovirus and adeno associated virus (AAV) expressing Cre at P1. Of all tested Cre-expressing viruses, LV-Cre showed the best labeling of the NSC in the SVZ and GFP-linage tracing of their progeny in the midbrain.
(**D**) Representative histological IHC images using an IDH1$^{R132H}$-specific antibody of an *Idh1*$^{+/+}$ (left) and *Idh1*$^{LSL-R132H/+}$ (right) brain transduced with LV- Cre. Scale bar, 100μm.
(**E**) 2HG levels in LSL-*Idh1*$^{R132H}$ mouse brains transduced with Ad-GFP and Ad-Cre-GFP. Statistical comparison was performed using two-tailed t-test.

**Fig. S5.** ***In vivo*** **CRISPR methodology in the Mouse Brain**

(**A**) Schematic depicting our *in vivo* double fluorescent reporter strategy to quantify efficacy of CRIPSR/Cas9-mutagenesis. LSL-Cas9-GFP; LSL-tdTomato mice transduced with LV-Cre encoding a scrambled sgRNA (LV-Scr-Cre) co-express green fluorescent protein (GFP) and tdTomato. Mice transduced with LV-Cre targeting GFP (LV-sgGFP-Cre) exhibit tdTomato$^{+}$ cells lacking GFP, allowing for quantification of GFP knockout efficacy by IF and/or FACS, revealing a knock-out efficiency of 85±5%.

(**B**) Representative fluorescent images of tdTomato and GFP expression within brains of LSL-Cas9-GFP TdTomato mice transduced with LV-Cre carrying either sgScr (top) or sgGFP (middle, bottom). White arrows indicate loss of GFP expression in tdTomato positive cells.

(**C**) Expression of GFP and tdTomato in brains of LSL-Cas9-GFP; LSL-tdTomato mice transduced with LV-Cre constructs expressing either sgScr (top left) or sgGFP (top right). Co-localization analysis and 10x magnification of respective images (bottom).

(**D**) Fluorescent image of a sagittal section of whole LSL-Cas9-GFP brain transduced with LV-sg*Urod*-Cre. To confirm efficient mutagenesis of an endogenous gene, we targeted the heme biosynthesis gene *Urod*. Loss of *Urod* leads to accumulation of unprocessed, fluorescent porphyrins (*73*) and bright red fluorescence in the SVZ, the cerebrum and olfactory bulb (OB)

(**E**) Bar graph showing spectrum of indels and their frequencies for two sgRNAs targeting *Atrx* using Tracking of Indels by Decomposition algorithm (TIDE).

**Fig. S6. Mouse model of LGG histologically recapitulates human tumors**

(**A**) Representative H&E and Ki67 staining of a tumor, a hyperplastic lesion and a lesion with increased proliferation in the subventricular zone (SVZ) illustrating the classification scheme used to determine cohort phenotypes in Fig. 3C and 4B. Scale bars, 2.5mm (top and middle) and 100μm (bottom).

(**B**) Representative brightfield and fluorescent image of mouse LGG brain (left) as well as H&E and IHC staining of the same tumor region within a sg*Atrx; Idh1*$^{R132H/+}$; *Trp53*$^{R270H/+}$; Cas9-GFP brain using the indicated antibodies. Scale bars, 2.5mm (left) and 50μm (right).

(**C**) Representative brightfield and fluorescent image of mouse LGG brain (top left) as well as H&E staining (top right), and immunohistochemical staining of the same tumor region within a sg*Atrx; Idh1*$^{R132H/+}$;*Trp53*$^{R270H/fl}$; Cas9-GFP brain using the indicated antibodies (bottom). Scale bars, 2.5mm for whole brain sections and 50μm for close-ups.

**Fig. S7. Mouse model of LGG molecularly recapitulates human tumors**
(**A**) Comparison of GSEA analysis of 50 Hallmark gene sets in mouse LGG vs. mouse brain. Bar graph shows the top differential pathways ranked by NES with nominal p-value <0.05 and GSEA plots show selected pathways.
(**B**) Unsupervised hierarchical clustering of RNA expression of mouse sg*Atrx*; *Idh1*$^{R132H/+}$; *Trp53*$^{R270H/+}$; Cas9-GFP brain tumors, human LGG and human gliosis.
(**C**) Representative whole brain fluorescent image, histological H&E staining and IHC for Ki67 of a brain injected with rs557$^{66bp/+}$,*Idh1*$^{R132H/+}$;*Trp53*$^{\Delta/\Delta}$;Cas9-GFP RIP cells. Scale bar, 100μm.

**Fig. S8. Generation of rs55705857-mimetic mice**

(**A**) Sequence alignment of human rs55705857 region on chromosome 8q.24.21 with the syntenic region on mouse chromosome 15.

(**B**) OCT2 (POU2F2) motif (top) aligned with the rs55705857 non-risk (middle) and risk (bottom) allele.

(**C**) Sanger sequencing results of the PCR amplicon encompassing the syntenic mouse rs55705857 region in wildtype and CRSIPR/Cas9-edited rs557$^{A\text{->}G+4}$ and rs557$^{66bp}$ mouse strains.

(**D**) Enrichment of H3K27Ac, H3K4me$^{1}$ and pan-H3 at mouse rs55705857 locus as determined by ChIP-qPCR using rs557$^{66bp/+}$, *Idh1*$^{R132H/+}$; *Trp53*$^{\Delta/\Delta}$-mutant RIP tumors cells (n = 3). IgG-IP serves as a negative control. Statistical comparison was performed using two-tailed t-test.

(**E**) Representative IHC staining of OCT2 and OCT4 expression within rs557$^{66bp/+}$, *Idh1*$^{R132H/+}$; *Trp53*$^{fl/fl}$; Cas9-GFP mouse LGG. Scale bars, 25µm.

(**F**) RNAseq results for *OCT4* (*POU5F1*), *OCT2* (*POU2F2*), *ASCL1, ASCL2* and *SOX2* from patient glioma and gliosis samples. Tumors are stratified by *IDH* mutation, significant comparisons are shown with their p-values. Non-significant comparisons are not shown.

(**G**) Western Blot analysis of OCT2 and OCT4 in cultured RIP cells. GAPDH serves as loading control.

(**H**) Bar graph showing spectrum of indels and their frequencies for a sgRNA targeting the WT rs55705857 allele in rs557$^{66bp/+}$ RIP cells using Tracking of indels by Decomposition algorithm (TIDE).

**Fig. S9. rs55705857 interacts with MYC promoter and modulates epigenetic landscape of 8q24**
Myc-FAM49b (Chr8:127,700,000-130,045,000; hg38) ATAC-seq data for the *IDH*-WT (8 GBM and 1 LGG) and *IDH*-mutant (12 LGG and 1 GBM) brain tumors and skin cutaneous melanoma (SKCM) are aligned with H3K27ac as well as DiffBind log2-fold change for H3K27ac and H3K4me$^1$ when comparing *IDH*-mutant vs *IDH*-WT brain tumors. ChIPseq signals for H3K27ac (dark green) are shown for the Mayo cohort of cases, stratified into *IDH*-WT, *IDH*-mutant non-codel and *IDH*-mutant codel groups. DiffBind shows the fold change and p-value between the *IDH*-mutant vs. *IDH*-WT for both H3K27c and H3K4me$^1$. ChromHMM shows the predicted function of the genome surrounding rs55705857 based on the histone marks H3K36me$^3$, H3K4me$^1$, H3K27ac and H3K4me$^3$ in *IDH*-WT and *IDH*-mutant brain tumors as well as non-tumorous gliosis samples. Interactions from Hi-C using the rs55705857 as the anchor point in hippocampus (Yang D et al, 2018) and Promoter-capture Hi-C interactions centered at the *MYC* promoter (*32*) are shown. The light blue line marks the location of rs55705857, and shaded gray box marks the location of *MYC*. *IDH*-mutant brain tumors and gliosis samples are sorted by rs55705857 non-risk (A) and risk (G) alleles.
