## Supplemental Figures for "A non-coding single nucleotide polymorphism at 8q24 drives IDH1-mutant glioma formation"

### Supplemental figures and figure legends

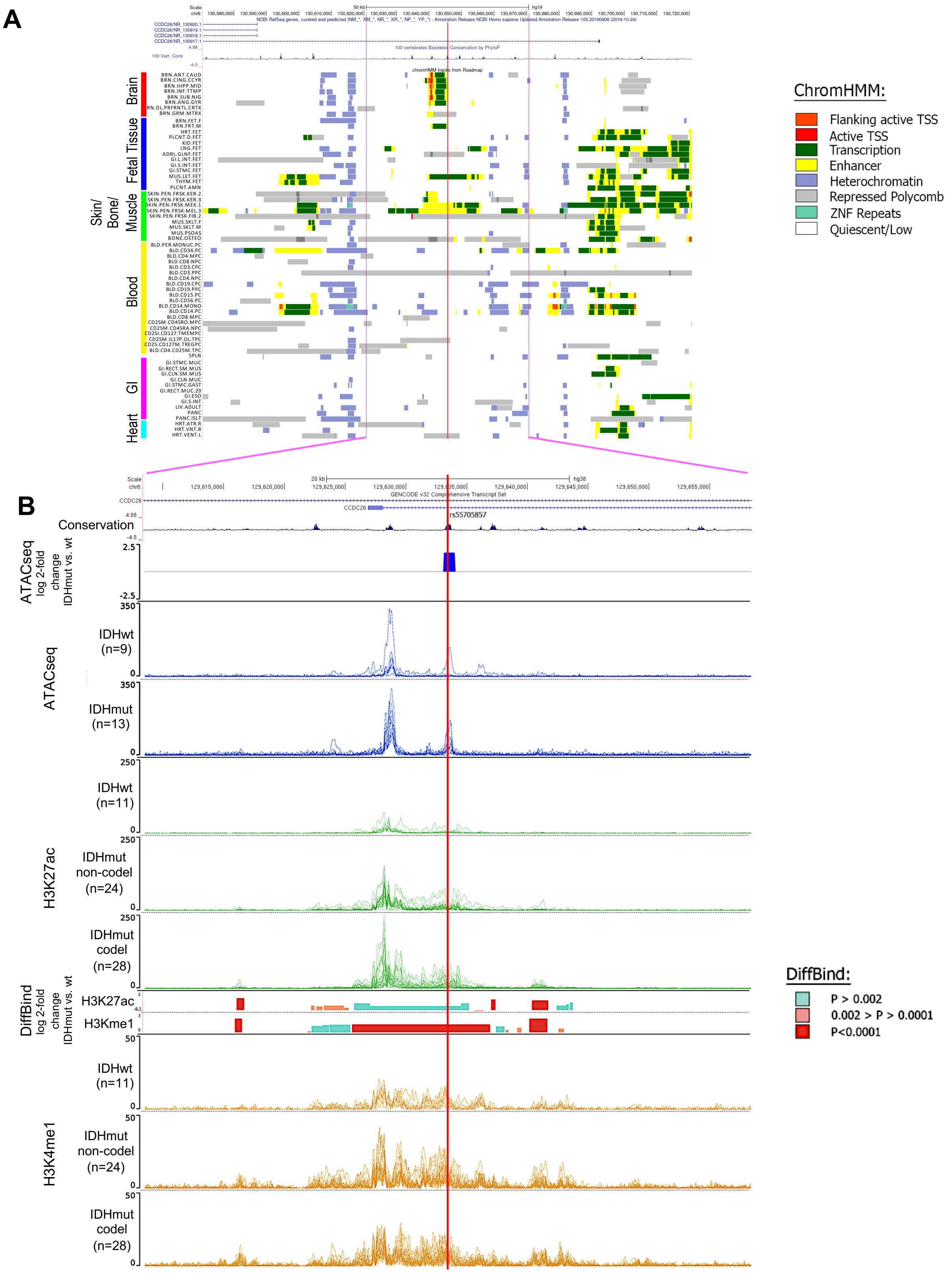


**Figure S1. rs55705857 is located in a brain-specific enhancer**

**Fig. S1. rs55705857 is located in a brain-specific enhancer at 8q24**

(**A**) Roadmap and ENCODE data showing the overlap between rs55705857 (indicated by the red line) and enhancer features in 64 normal tissues and normal blood, including the brain.

(**B**) Genomic locus surrounding rs55705857 (chr8:129,608,396-129,658,495; hg38). ATACseq data from the TCGA LGG and GBM, stratified by *IDH* mutation status, are displayed, each line represents an individual tumor (dark blue tracings). The fold change between the ATACseq signal for *IDH*-mutant vs. *IDH*-WT is displayed just above the tracings (blue, 0.001>p>0.002). ChIPseq signals for H3K27ac (dark green) and H3K4me[1] (orange) are shown for the Mayo cohort of tumors, stratified into *IDH*-WT, *IDH*-mutant non-codel and *IDH*-mutant codel groups. DiffBind shows the fold change and p-value between the *IDH*-mutant vs. *IDH*-WT for both H3K27c and H3K4me[1] (blue, 0.001>P>0.002; pink, 0.002>P>0.0001; red, P<0.0001). Red line indicates position of rs55705857.

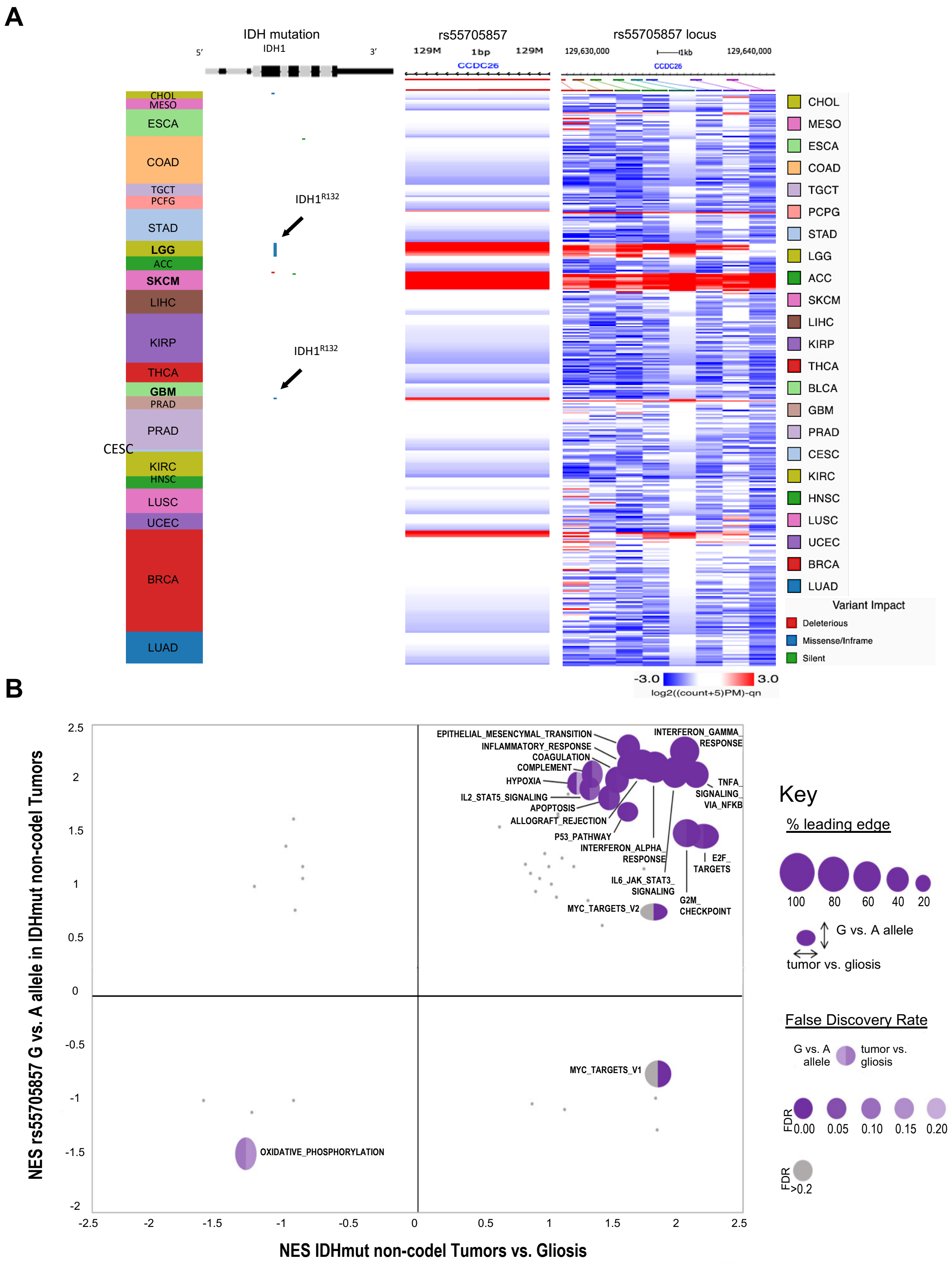


**Fig. S2. rs55705857 risk allele G enhances an LGG-transcriptional profile**
(**A**) *IDH1* mutation and ATAC-seq accessibility at rs55705857 SNP (chr8:129,633,446; hg38) and at genomic locus surrounding rs55705857 (chr8:129,630,000-129,640,000) in 377 TCGA samples across 23 cancer types using UCSC Xena. (**B**) Comparison of GSEA results using 50 Hallmark gene sets in *IDH*-mutant non-codel tumors vs. gliosis and rs55705857 A vs. G allele in *IDH*-mutant non-codel tumors.

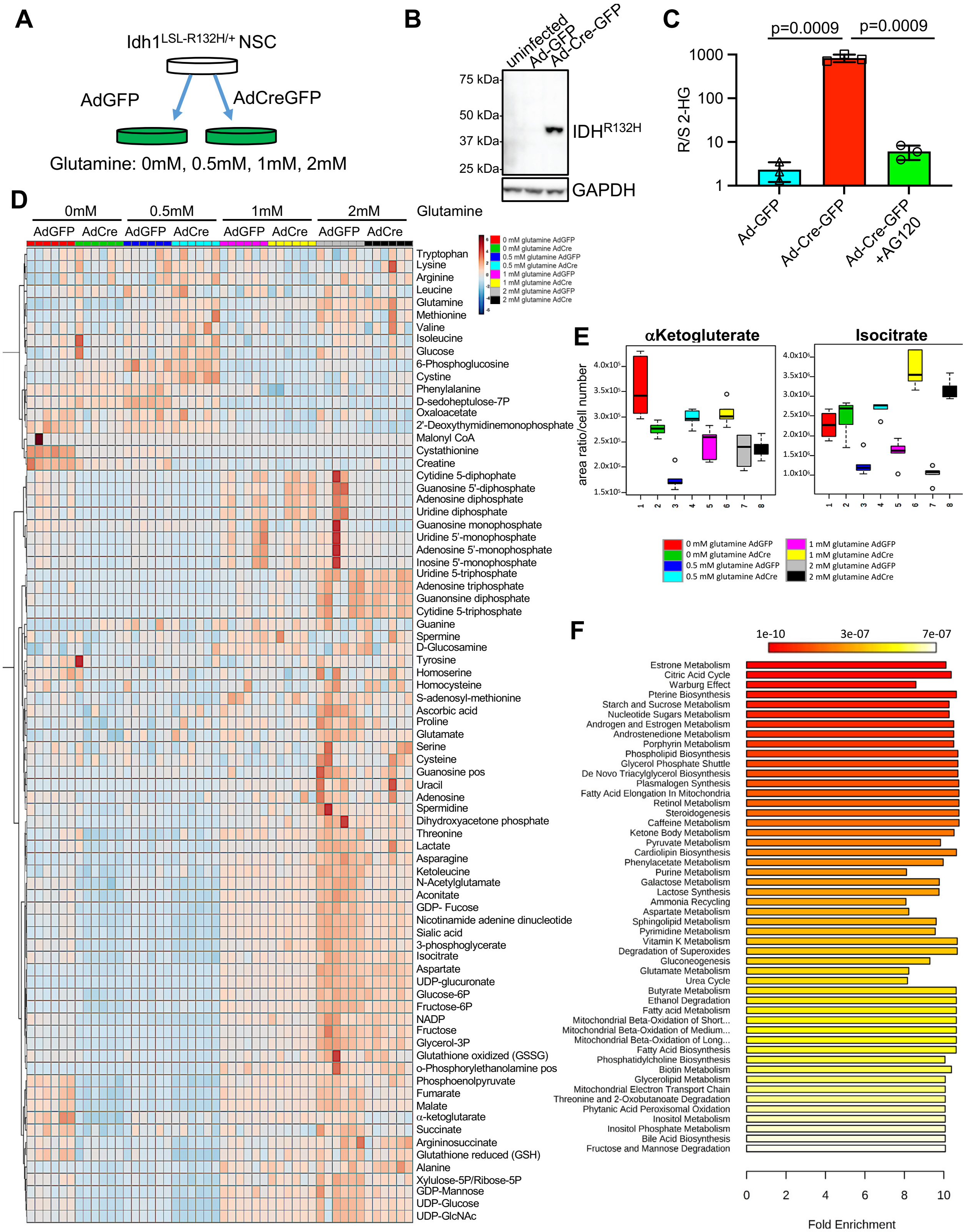


**Figure S3. IDH1$^{R132H}$ metabolically rewires NSCs**

**Fig. S3. IDH1$^{R132H}$ metabolically rewires NSC**
(**A**) Schematic of IDH1$^{R132H}$ induction in mouse NSC.
(B) Western blot of IDH1$^{R132H}$ expression in uninfected or Ad-GFP or Ad-Cre-GFP transduced LSL-*Idh1*$^{R132H}$ mouse NSC using an IDH1$^{R132H}$ specific antibody.
(**C**) Ratio of R- to S-2HG in LSL-*Idh1*$^{R132H}$ mouse NSC transduced with Ad-GFP or Ad-Cre-GFP and treated with or without AG120 measured by mass spectrometry. Statistical comparison was performed using two-tailed t-test.
(**D**) Heatmap of 79 metabolites in syngeneic *Idh1*$^{+/+}$ and *Idh1*$^{R132H/+}$ NSC cultured in 4 different glutamine concentrations measured in sextuplicate by mass spectrometry.
(**E**) Normalized concentrations of isocitrate and the α-ketoglutarate in syngeneic *Idh1*$^{+/+}$ and *Idh1*$^{R132H/+}$ NSC cultured in 4 different glutamine concentrations measured in sextuplicate.
(**F**) Bar chart of enrichment analysis of metabolic pathways altered in syngeneic *Idh1*$^{R132H/+}$ compared to syngenic *Idh1*$^{+/+}$ NSC cultured in 4 different glutamine concentrations. Scale bar at the top indicate p-value.

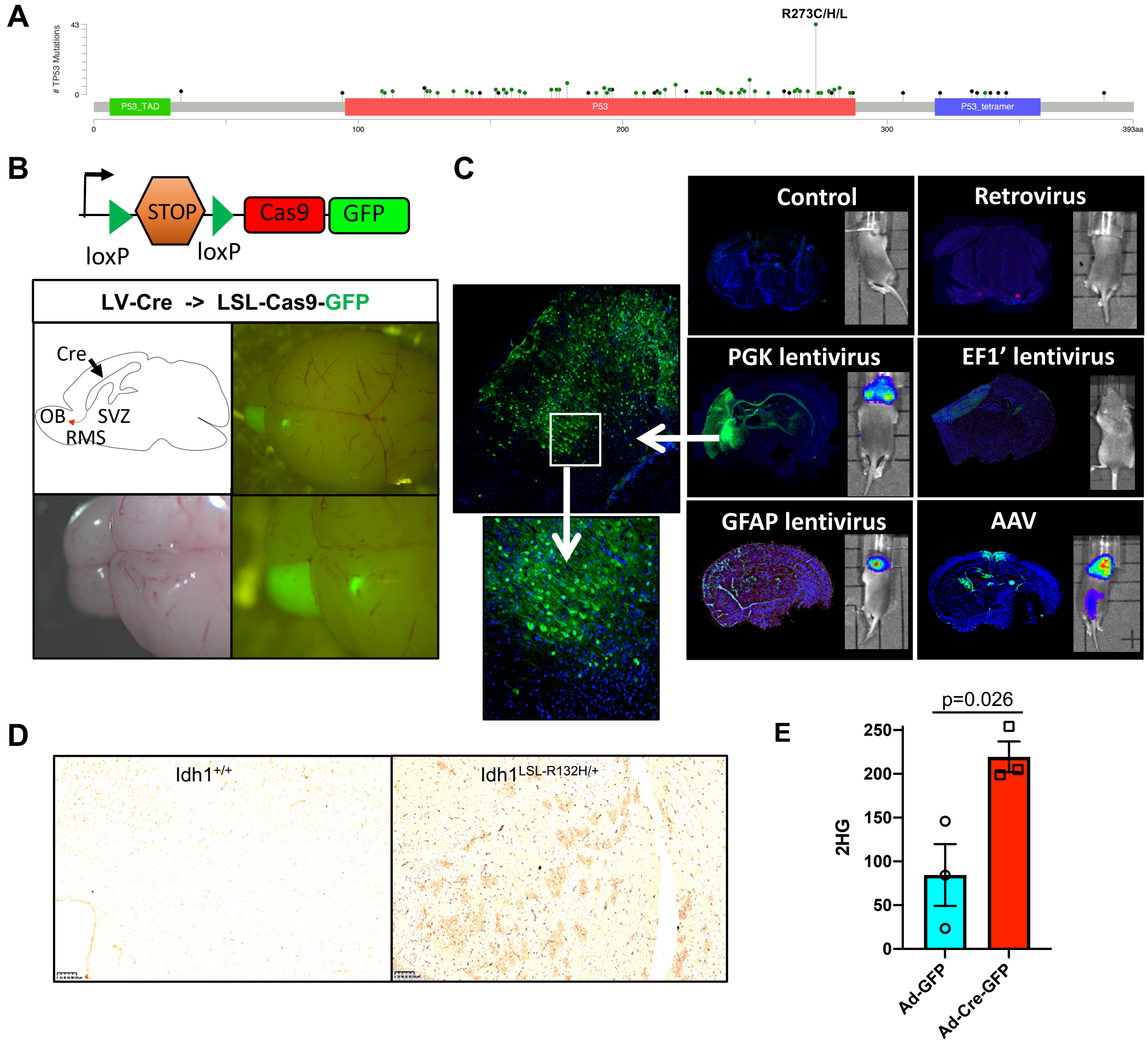


**Fig. S4.** ***In vivo*** **Transduction of the Mouse Brain**

(**A**) Lollipop plot depicting *TP53* mutations found in human LGG (data from the TCGA database).

(**B**) Schematic of the LSL-Cas9-GFP mouse cassette (Top). Schematic of the viral Cre injection targeting NSC of the SVZ of LSL-Cas9-GFP reporter mice, which leads to linage tracing of neural stem/progenitors and their progeny in the rostral migratory stream (RMS) and olfactory bulb (OB). Brightfield and fluorescent images of LSL-Cas9-GFP mouse brains at postnatal day 21 (P21) transduced with lentiviral Cre at P1.

(**C**) Representative immunofluorescent and bioluminescence images of LSL-Cas9-GFP brains at postnatal day 7 (P7) transduced with lentivirus (LV), adenovirus and adeno associated virus (AAV) expressing Cre at P1. Of all tested Cre-expressing viruses, LV-Cre showed the best labeling of the NSC in the SVZ and GFP-linage tracing of their progeny in the midbrain.

(**D**) Representative histological IHC images using an IDH1$^{R132H}$-specific antibody of an *Idh1*$^{+/+}$ (left) and *Idh1*$^{LSL\text{-}R132H/+}$ (right) brain transduced with LV- Cre. Scale bar, 100µm.

(**E**) 2HG levels in LSL-*Idh1*$^{R132H}$ mouse brains transduced with Ad-GFP and Ad-Cre-GFP. Statistical comparison was performed using two-tailed t-test.

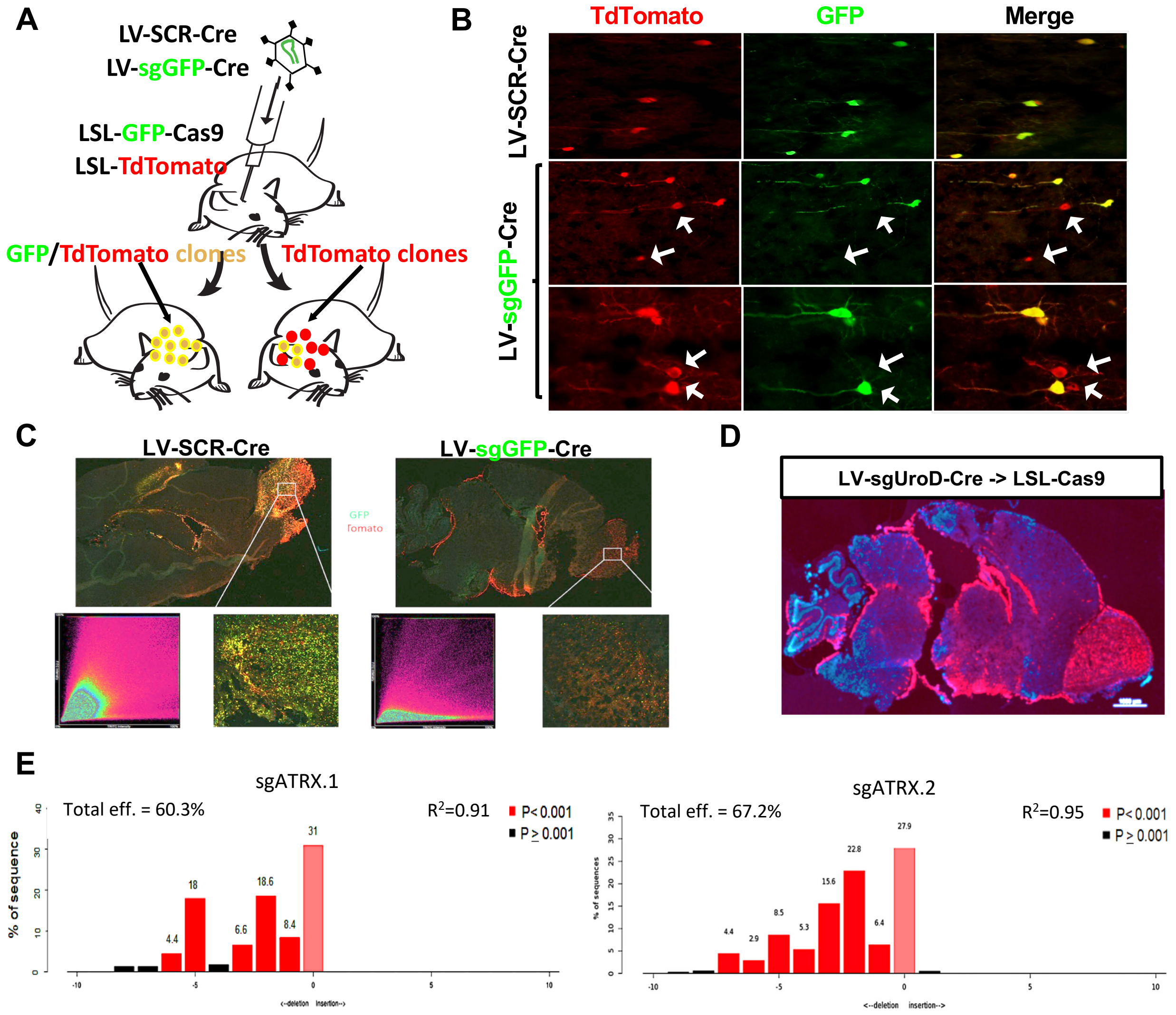


**Fig. S5.** ***In vivo*** **CRISPR methodology in the Mouse Brain**

(**A**) Schematic depicting our *in vivo* double fluorescent reporter strategy to quantify efficacy of CRIPSR/Cas9-mutagenesis. LSL-Cas9-GFP; LSL-tdTomato mice transduced with LV-Cre encoding a scrambled sgRNA (LV-Scr-Cre) co-express green fluorescent protein (GFP) and tdTomato. Mice transduced with LV-Cre targeting GFP (LV-sgGFP-Cre) exhibit tdTomato$^+$ cells lacking GFP, allowing for quantification of GFP knockout efficacy by IF and/or FACS, revealing a knock-out efficiency of 85±5%.

(**B**) Representative fluorescent images of tdTomato and GFP expression within brains of LSL-Cas9-GFP TdTomato mice transduced with LV-Cre carrying either sgScr (top) or sgGFP (middle, bottom). White arrows indicate loss of GFP expression in tdTomato positive cells.

(**C**) Expression of GFP and tdTomato in brains of LSL-Cas9-GFP; LSL-tdTomato mice transduced with LV-Cre constructs expressing either sgScr (top left) or sgGFP (top right). Co-localization analysis and 10x magnification of respective images (bottom).

(**D**) Fluorescent image of a sagittal section of whole LSL-Cas9-GFP brain transduced with LV-sg*Urod*-Cre. To confirm efficient mutagenesis of an endogenous gene, we targeted the heme biosynthesis gene *Urod*. Loss of *Urod* leads to accumulation of unprocessed, fluorescent porphyrins (*73*) and bright red fluorescence in the SVZ, the cerebrum and olfactory bulb (OB)

(**E**) Bar graph showing spectrum of indels and their frequencies for two sgRNAs targeting *Atrx* using Tracking of Indels by Decomposition algorithm (TIDE).

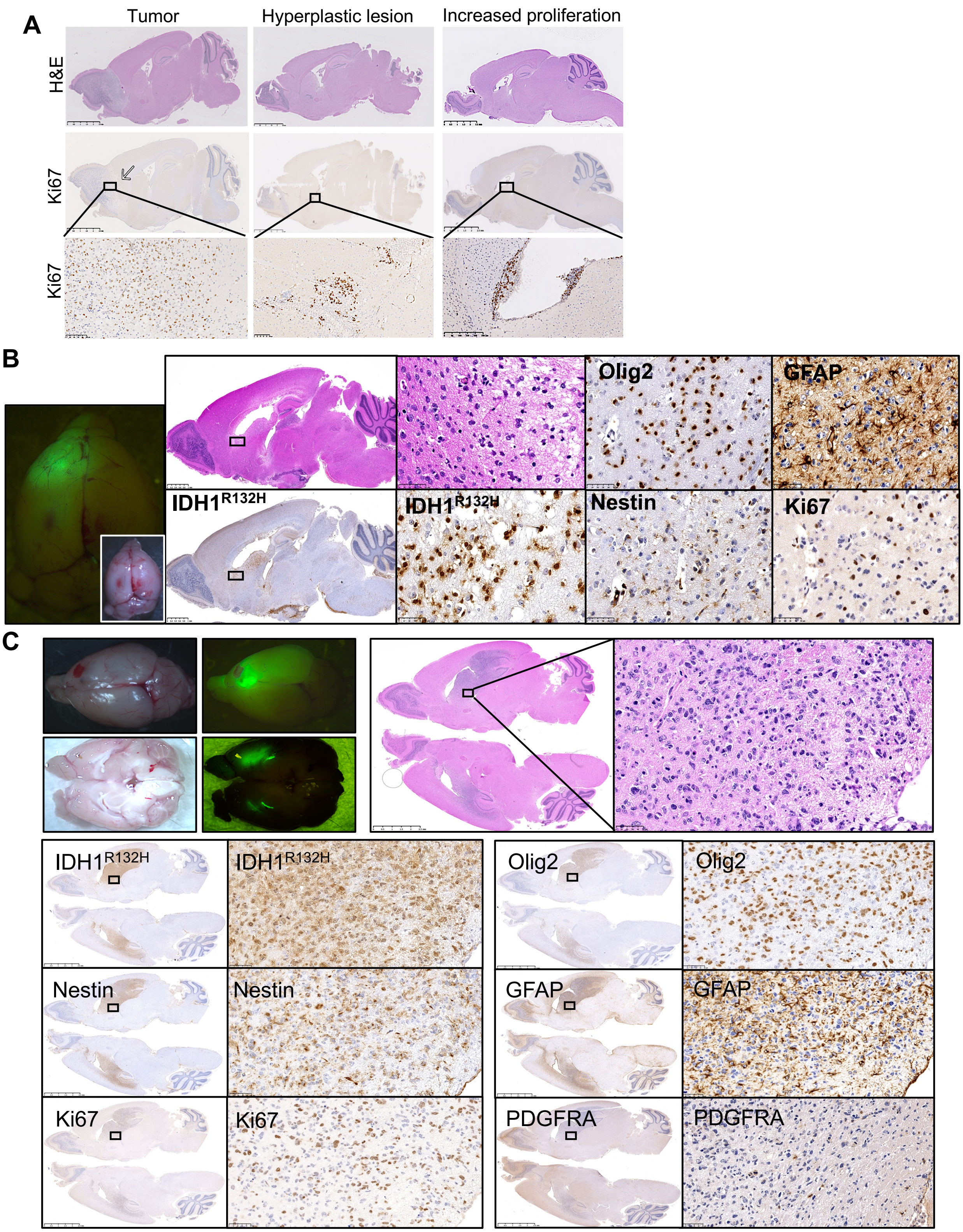


**Figure S6. Mouse model phenotypically recapitulates human LGGs**

**Fig. S6. Mouse model of LGG histologically recapitulates human tumors**

(**A**) Representative H&E and Ki67 staining of a tumor, a hyperplastic lesion and a lesion with increased proliferation in the subventricular zone (SVZ) illustrating the classification scheme used to determine cohort phenotypes in Fig. 3C and 4B. Scale bars, 2.5mm (top and middle) and 100µm (bottom).

(**B**) Representative brightfield and fluorescent image of mouse LGG brain (left) as well as H&E and IHC staining of the same tumor region within a sg*Atrx; Idh1*$^{R132H/+}$; *Trp53*$^{R270H/+}$; Cas9-GFP brain using the indicated antibodies. Scale bars, 2.5mm (left) and 50µm (right).

(**C**) Representative brightfield and fluorescent image of mouse LGG brain (top left) as well as H&E staining (top right), and immunohistochemical staining of the same tumor region within a sg*Atrx; Idh1*$^{R132H/+}$;*Trp53*$^{R270H/fl}$; Cas9-GFP brain using the indicated antibodies (bottom). Scale bars, 2.5mm for whole brain sections and 50µm for close-ups.

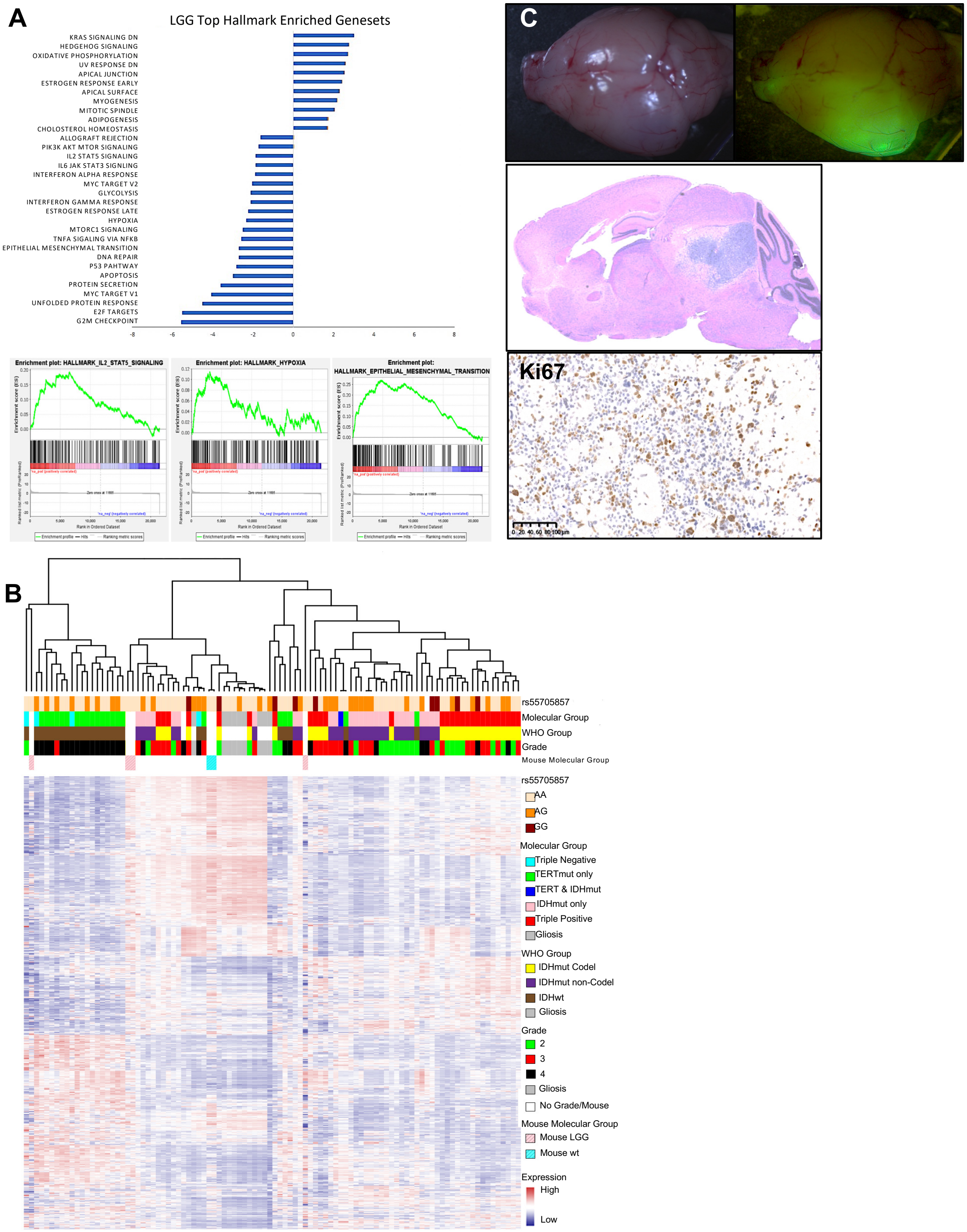


**Figure S7. Mouse model phenotypically recapitulates human LGGs**

**Fig. S7. Mouse model of LGG molecularly recapitulates human tumors**
(**A**) Comparison of GSEA analysis of 50 Hallmark gene sets in mouse LGG vs. mouse brain. Bar graph shows the top differential pathways ranked by NES with nominal p-value <0.05 and GSEA plots show selected pathways.
(**B**) Unsupervised hierarchical clustering of RNA expression of mouse sg*Atrx*; *Idh1*$^{R132H/+}$; *Trp53*$^{R270H/+}$; Cas9-GFP brain tumors, human LGG and human gliosis.
(**C**) Representative whole brain fluorescent image, histological H&E staining and IHC for Ki67 of a brain injected with rs557$^{66bp/+}$,*Idh1*$^{R132H/+}$;*Trp53*$^{\Delta/\Delta}$;Cas9-GFP RIP cells. Scale bar, 100µm.

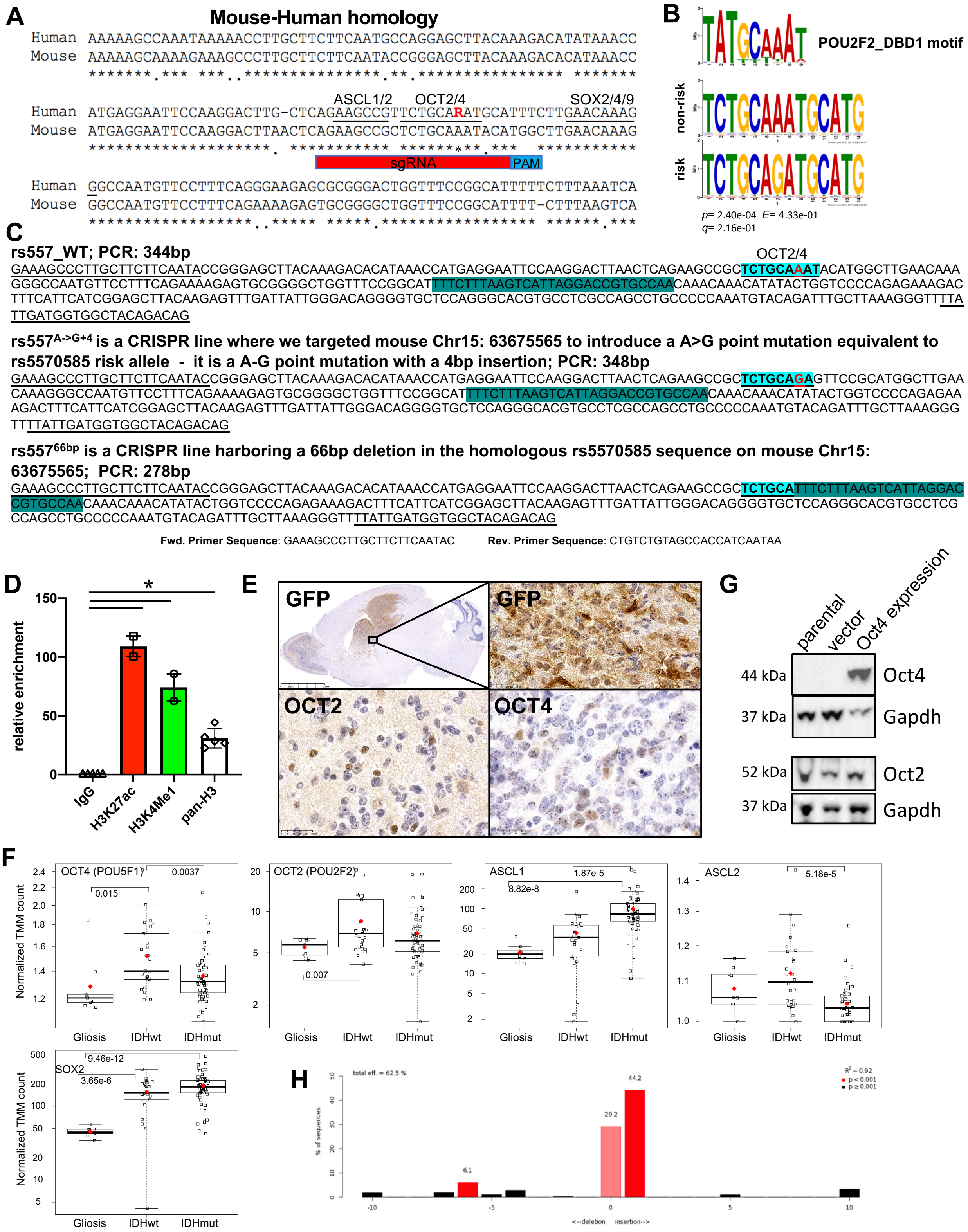


### Figure S8. Generation of rs55705857-mimetic mice

**Fig. S8. Generation of rs55705857-mimetic mice**

(**A**) Sequence alignment of human rs55705857 region on chromosome 8q.24.21 with the syntenic region on mouse chromosome 15.

(**B**) OCT2 (POU2F2) motif (top) aligned with the rs55705857 non-risk (middle) and risk (bottom) allele.

(**C**) Sanger sequencing results of the PCR amplicon encompassing the syntenic mouse rs55705857 region in wildtype and CRSIPR/Cas9-edited rs557$^{A\text{->}G+4}$ and rs557$^{66bp}$ mouse strains.

(**D**) Enrichment of H3K27Ac, H3K4me$^{1}$ and pan-H3 at mouse rs55705857 locus as determined by ChIP-qPCR using rs557$^{66bp/+}$, *Idh1*$^{R132H/+}$; *Trp53*$^{\Delta/\Delta}$-mutant RIP tumors cells (n = 3). IgG-IP serves as a negative control. Statistical comparison was performed using two-tailed t-test.

(**E**) Representative IHC staining of OCT2 and OCT4 expression within rs557$^{66bp/+}$, *Idh1*$^{R132H/+}$; *Trp53*$^{fl/fl}$; Cas9-GFP mouse LGG. Scale bars, 25µm.

(**F**) RNAseq results for *OCT4* (*POU5F1*), *OCT2* (*POU2F2*), *ASCL1, ASCL2* and *SOX2* from patient glioma and gliosis samples. Tumors are stratified by *IDH* mutation, significant comparisons are shown with their p-values. Non-significant comparisons are not shown.

(**G**) Western Blot analysis of OCT2 and OCT4 in cultured RIP cells. GAPDH serves as loading control.

(**H**) Bar graph showing spectrum of indels and their frequencies for a sgRNA targeting the WT rs55705857 allele in rs557$^{66bp/+}$ RIP cells using Tracking of indels by Decomposition algorithm (TIDE).

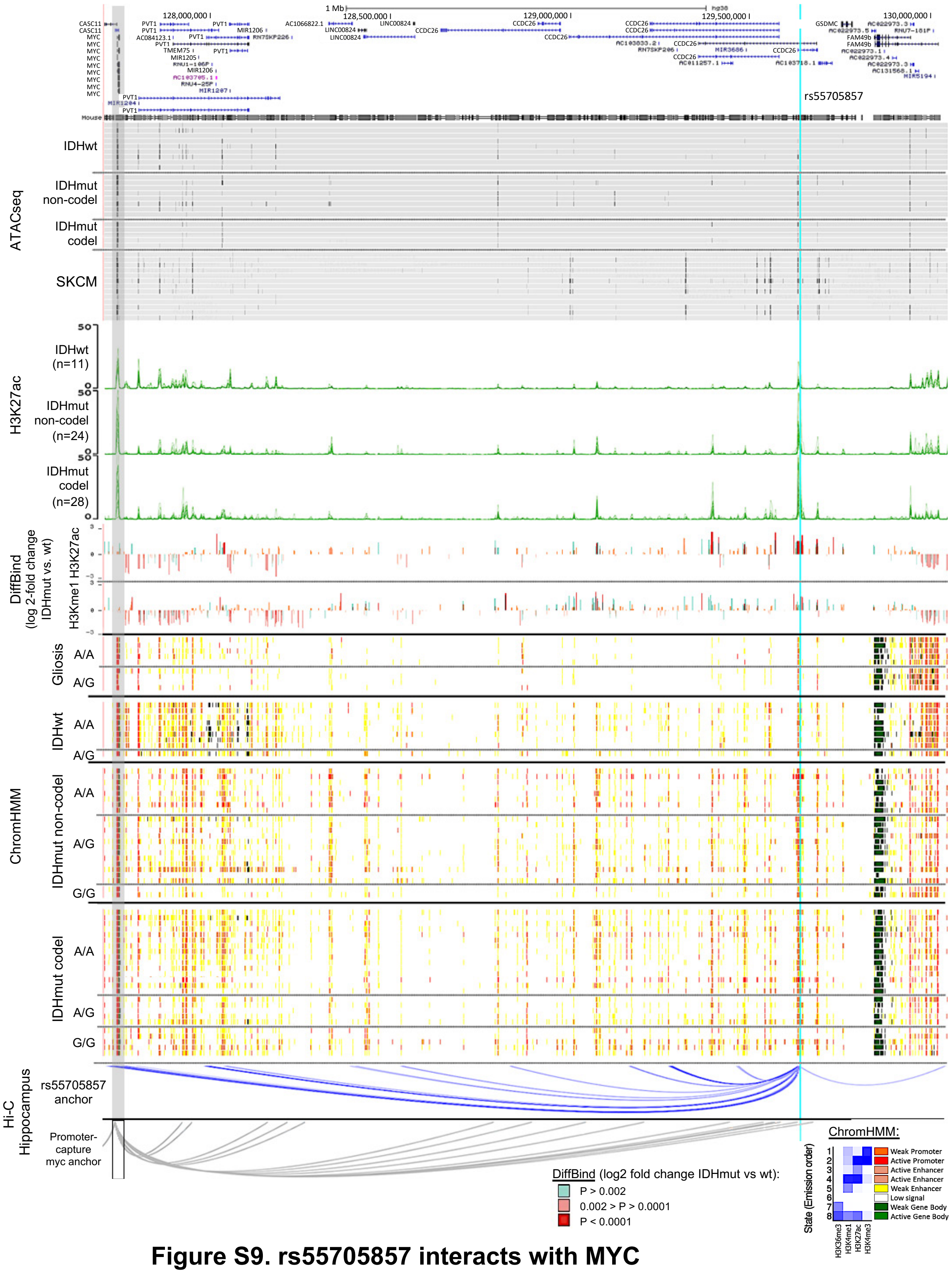


**Figure S9. rs55705857 interacts with MYC**

**Fig. S9. rs55705857 interacts with MYC promoter and modulates epigenetic landscape of 8q24**
Myc-FAM49b (Chr8:127,700,000-130,045,000; hg38) ATAC-seq data for the *IDH*-WT (8 GBM and 1 LGG) and *IDH*-mutant (12 LGG and 1 GBM) brain tumors and skin cutaneous melanoma (SKCM) are aligned with H3K27ac as well as DiffBind log2-fold change for H3K27ac and H3K4me$^1$ when comparing *IDH*-mutant vs *IDH*-WT brain tumors. ChIPseq signals for H3K27ac (dark green) are shown for the Mayo cohort of cases, stratified into *IDH*-WT, *IDH*-mutant non-codel and *IDH*-mutant codel groups. DiffBind shows the fold change and p-value between the *IDH*-mutant vs. *IDH*-WT for both H3K27c and H3K4me$^1$. ChromHMM shows the predicted function of the genome surrounding rs55705857 based on the histone marks H3K36me$^3$, H3K4me$^1$, H3K27ac and H3K4me$^3$ in *IDH*-WT and *IDH*-mutant brain tumors as well as non-tumorous gliosis samples. Interactions from Hi-C using the rs55705857 as the anchor point in hippocampus (Yang D et al, 2018) and Promoter-capture Hi-C interactions centered at the *MYC* promoter (*32*) are shown. The light blue line marks the location of rs55705857, and shaded gray box marks the location of *MYC*. *IDH*-mutant brain tumors and gliosis samples are sorted by rs55705857 non-risk (A) and risk (G) alleles.
